# Unverified Vendor Claims and Preventable Harms: A Mixed-Methods Longitudinal Independent Audit of Health AI System Performance in Nigeria

**DOI:** 10.64898/2026.03.21.26348981

**Authors:** Benjamin S. Chudi Uzochukwu, Yakubu Joel Cherima, Ugo Uwadiako Enebeli, Blessing Hassan, Chinyere Cecelia Okeke, Adaora Chinelo Uzochukwu, Amobi Omoha, Kennedy Anenechukwu Uzochukwu, Eziyi Iche Kalu, Daniel Victor, Habibatu Excellent Alih, Ladi Sule Matinja, Innocent Timothy Rindap

## Abstract

**Objective:** To independently audit vendor-reported performance claims of health AI systems deployed in Nigeria and assess discrepancies, clinical consequences, equity impacts, and implications for safe AI deployment in low- and middle-income countries.

**Methods and analysis:** We conducted a mixed-methods longitudinal audit (October 2024-March 2026) of six health AI systems (chest X-ray interpretation, TB screening, symptom triage, maternal health risk prediction, patient history intake, and health chatbots) across 73 diverse health facilities in six Nigerian states, involving 52,000 patients and 45 key informant interviews conducted with stakeholders. All data were sourced from integrated facility-level records, and no database linkage was performed. Vendor claims were abstracted from documentation, white papers, and validation studies. Independent performance was verified by an independent third party through system logs, patient records, clinical outcomes, and stakeholder interviews. Performance gaps were quantified as absolute percentage-point differences; clinical harms were estimated using patient volume and bootstrap confidence intervals; equity impacts were assessed across vulnerability dimensions (geography, age, income, comorbidities, infrastructure) using interaction terms in mixed-effects models and an Equity Harm Index (EHI).

**Results:** Vendor-reported accuracy averaged 91.5%, while independently measured real-world accuracy averaged 67.3%, yielding a mean performance gap of 24.2 percentage points (95% CI: 21.5 to 26.9; p<0.001) across systems. Gaps ranged from 17 to 35 percentage points and were statistically significant for all systems. These discrepancies translated to substantial preventable harm, including an estimated 1,247 undetected TB cases (186 preventable deaths) and 342 misclassified high-risk pregnancies annually. Performance gaps were 28-38% larger among vulnerable groups (e.g., rural patients showed 38% higher EHI). Gaps were classified as systematic, context-dependent, or population-dependent.

**Conclusion:** Vendor-reported performance metrics substantially overstated the real-world effectiveness of health AI in Nigeria, leading to preventable patient harm and widening inequities. Mandatory independent post-deployment verification, analogous to pharmaceutical Phase IV surveillance, is essential to ensure safe, equitable AI use in resource-constrained settings. Donors and regulators should prioritize verification over trust-based deployment.

**KEY MESSAGES:** *What is already known on this topic:*

- Health AI systems are rapidly deployed in LMICs, mainly based on vendor-reported performance from controlled or retrospective validations, often accepted without independent real-world checks.
- Evidence suggests that discrepancies arise from domain shifts, data quality issues, and population differences, but large-scale, independent audits in LMIC settings are scarce, creating a ‘verification paradox’.

*What this study adds:*

- This audit of six health AI systems across 73 Nigerian facilities and 52,000 patients found a consistent 24.2 percentage-point gap (95% CI 21.5-26.9) between vendor-reported accuracy (mean 91.5%) and real-world performance (mean 67.3%), causing preventable harms including about 1,247 undetected TB cases, 186 preventable deaths, and 342 misclassified high-risk pregnancies annually.
- The gaps were 28-38% larger in vulnerable groups (e.g., 38% higher Equity Harm Index in rural patients) and classified as systematic, context-dependent, or population-dependent.
- How this study might affect research, practice or policy
- Vendor self-reporting is unreliable, and mandatory independent post-deployment verification, similar to pharmaceutical Phase IV surveillance, is essential for patient safety and equity in global health AI. Donors, governments, and regulators should adopt verification-based models to prevent harm, reduce inequalities, and inform future multinational studies and LMIC regulatory frameworks.

## 1. INTRODUCTION

As artificial intelligence (AI) tools are rapidly integrated into health systems across Low- and Middle-Income Countries (LMICs), the question of whether AI systems actually work after deployment has become increasingly urgent.^1,2^ AI-powered technologies are now regularly adopted by donors, governments, and non-governmental organisations, with the promise of improving the quality, effectiveness, and equity of care, from diagnostic decision support to patient groups and disease supervision.^3^ There is still a serious gap between how AI performance is reported before deployment and how it actually performs in real-world health systems.^4^ Recent procurement and financing decisions for health AI systems are heavily influenced by vendor-reported precision metrics, typically derived from controlled testing environments. These precision metrics are usually derived from retrospective validation, curated datasets, and pilot studies conducted in ideal conditions. Donors and implementing partners frequently accept such claims at face value, using them as primary evidence that a system is effective or ready for scale.^5^

In practice, little systematic effort is made to independently verify whether these reported performance levels persist after deployment, when AI tools encounter heterogeneous patient populations, variable clinical workflows, data quality issues, and infrastructure constraints.^6^ Dependence on vendor-reported performance highlights a serious scientific issue regarding accountability and credibility. AI vendors operate under transparent conflicts of interest, where funding strongly favours reporting optimistic results, while weak performance reduces revenue and future contracts.^7^ Favourable precision claims, renewals, continued sales, and donor endorsements are among the benefits for vendors that depend on perceived effectiveness. Conversely, biases that expose failures and performance degradation in real-world settings result in substantial commercial risk. It is therefore suggested that vendors should specifically report outputs, maximize evaluation contracts to increase metrics, or hinder disclosure of post-deployment limitations.^8^

In the pharmaceutical industry, accurate models that assess the risks of relying solely on manufacturer-reported efficacy are available. Drug companies usually magnify treatment benefits and underreport adverse effects before the widespread adoption of independent clinical trials and regulatory oversight. A meta-analysis revealed publication bias, with industry-funded trials more likely to report positive outcomes than independently funded studies.^9,10^ Independent verification has become the foundation of evidence-based medicine through regulatory audits, randomized controlled trials and post-marketing surveillance. Presently, there is no significant health system that can adopt a pharmaceutical effectiveness claim without external validation. The World Health Organization observed that many artificial intelligence (AI) methods are deployed without thorough evaluation.^1^ This is especially common in low- and middle-income countries (LMICs), where regulatory resources are often limited, and there is a strong push to quickly implement new technologies.

Consequently, the field faces the potential to establish a system in which vendors can readily validate their performance assertions, rather than subjecting them to independent testing under authentic operational conditions. Over $5 billion has been allocated to deploy health AI and digital health systems in LMICs with the objective of augmenting diagnostic capabilities and clinical decision support in environments with limited resources.^11,12^ Nevertheless, this swift expansion has outstripped the development of comprehensive assurance protocols, leading to a significant disparity between the scale of implementation and the robustness of regulatory oversight.

A critical paradox arises: the global health community is perhaps inadvertently cultivating a two-tiered system of AI safety, with countries with high incomes benefiting from established regulatory systems, such as those requiring FDA approval and independent validation. Thus, individuals in LMICs frequently encounter technologies that have not undergone thorough assessment, potentially exposing them to risks. Several factors contribute to this difference. Initially, companies frequently prioritize introducing their products in LMICs due to the comparatively less stringent regulatory environment. Secondly, LMICs often lack the requisite technical infrastructure for independent validation. In addition, donors and government agencies often prioritize quick implementation over thorough evaluation. This paper introduces a new way to classify performance gaps, one that looks beyond simple accuracy metrics. This classification will help with understanding the underlying causes of these gaps. Systematic disparities that affect all groups equally arise from fundamental differences between the environments in which the system was trained and those in which it is used. Context-dependent gaps, on the other hand, vary based on specific facility characteristics like infrastructure, workflow, and data quality and are caused by environmental factors.

Finally, population-dependent gaps, which disproportionately affect vulnerable groups such as the elderly, those in rural areas, and low-income individuals, stem from differences in data and algorithmic bias. For example, this study views independent verification not just as a technical task but as a key part of strengthening health systems. This study addresses four research questions:

1. How do vendor-reported performance claims compare to independently measured performance?
2. What are the clinical consequences of performance discrepancies?
3. Are these performance gaps evenly spread, or do they tend to impact more vulnerable groups?
4. Which framework is best for guiding independent verification in settings with limited resources?

## 2. METHODS

### 2.1 Study Design and Setting

A mixed-methods longitudinal audit was undertaken to assess the performance of AI within Nigerian healthcare institutions. Data collection was conducted between October 2024 and March 2026 at 73 public and private hospitals across six Nigerian states: Lagos, Gombe, Rivers, Enugu, Kaduna, and Plateau. The selected facilities represented a combination of urban (38) and rural (35) environments, thereby mirroring the infrastructural and operational diversity characteristic of low- and middle-income countries (LMICs). The primary objective of the audit was to evaluate the real-world efficacy of AI systems relative to vendor-supplied assertions, with the aim of identifying systemic performance deficiencies that could compromise patient safety.

### 2.2 Study Population and AI Systems

The study employed 52,000 participants across six AI systems deployed in clinical, diagnostic, and decision-making processes. The AI applications assessed were chest X-ray interpretation (Qure.ai qXR), Tuberculosis (TB) screening (Delft CAD4TB), symptom triage, maternal health risk prediction, health chatbots and patient history intake. Patients represented diverse demographic and linguistic backgrounds, including Nigerian English speakers, local languages, and Pidgin English.^13^ The combination of resource-constrained rural facilities and high-volume urban facilities provided awareness of the impact of patient population mix, infrastructure, and workflow inconsistencies on AI performance.

Inclusion into the study was determined by direct enumeration of patients whose interactions with the six AI systems occurred at participating facilities during the study period, with selection based on AI system deployment status and availability of corresponding system logs and patient records for independent verification. Consequently, no codes or algorithms were used to select the study population, and validation studies of selection codes or algorithms were not applicable.

### 2.3 Vendor Claim Abstraction and Verification

A comprehensive examination of vendor documentation was undertaken, encompassing: (1) white papers and technical documents; (2) marketing materials and promotional claims; (3) published validation studies; (4) regulatory submissions; and (5) internal documents furnished to healthcare facilities. The following information were collected for each AI system: (a) the main performance measure used (like sensitivity, specificity, or accuracy); (b) the dataset used for validation; (c) the characteristics of the study population; (d) the clinical setting; and (e) the reported performance results, including confidence intervals if they were available. A consistent method was utilized to extract vendor claims: (1) Two independent reviewers extracted claims from each document; (2) any disagreements were resolved through discussion or by reviewing the original document; (3) claims were categorized by type (sensitivity, specificity, accuracy, etc.); and (4) claims were evaluated for the quality of evidence using a 5-point scale, where 1 meant it was a marketing claim only, and 5 meant it was from a peer-reviewed publication with detailed methods. When multiple claims were found for the same system, the following was carried out: (1) identification of the most recent claim; (2) identification of claims from peer-reviewed publications (prioritized over marketing materials); (3) calculation of the mean and range of claims; (4) documentation of discrepancies between sources. For the TB screening system, the claims ranged from 92% to 96% accuracy across different vendor documents, and the most conservative estimate (92%) was used for comparison to independently measure performance.

### 2.4 Clinical Consequence Analysis

For each AI system, the key adverse outcomes associated with performance gaps were identified: (1) TB screening, missed TB cases (false negatives); (2) Maternal health, misclassified pregnancies (false positives), missed high-risk pregnancies (false negatives); (3) Malaria diagnosis, missed cases, unnecessary treatment. For each adverse outcome, there was (a) identification of cases from facility records; (b) linkage to AI system performance using causal inference methods; and (c) calculation of the rate and 95% CI.

For each adverse outcome, the following were calculated:

1. Number of cases attributable to performance gap = Performance Gap × Patient Volume;
2. Annual rate = Number of cases / 12 months; and
3. 95% CI using bootstrap resampling (n=10,000 iterations).

The difference in tuberculosis screening performance was 24 percentage points. With an annual patient volume of 50,000, this meant 12,000 cases were missed each year (0.24 × 50,000). Consequently, the clinical significance of these quantifiable harms was assessed: (1) For tuberculosis, the effect on mortality was estimated by multiplying the TB mortality rate by the number of missed cases; (2) For maternal health, the negative outcomes for mothers and newborns were estimated; (3) For malaria, the impact on both illness and death was estimated. And consultations were held with clinical experts to ensure interpretations were clinically meaningful.

### 2.5 Equity Analysis Methodology

Vulnerable populations were defined across five dimensions: (1) Demographic; age extremes (>65 years vs. <65 years); (2) Socioeconomic; income quintile (lowest vs. highest); (3) Geographic; location (rural vs. urban); (4) Clinical; presence of comorbidities (yes vs. (5) Infrastructure; the capacity of facilities, which can be classified as either low-capacity or high-capacity. For each dimension, the performance differences were compared between the privileged and vulnerable groups. Logistic regression with interaction terms was utilized to analyze the data, separating it by vulnerability dimension:

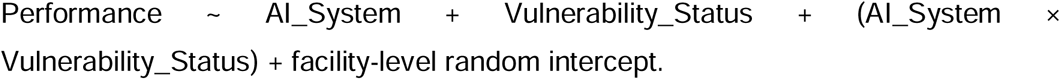

The interaction term was used to see if the performance differences were significantly larger for the vulnerable groups. The 95% confidence intervals were calculated using bootstrap resampling, with 10,000 iterations. To assess the unequal impact of performance gaps on vulnerable groups, an Equity Harm Index (EHI) was created:

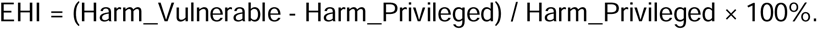

A positive Environmental Health Index (EHI) score demonstrated a disproportionate impact on vulnerable populations. In rural areas, patients experienced 38% more negative effects from tuberculosis screening compared to those in cities (EHI = 38%).

The mechanisms driving differential performance gaps were analyzed by examining:

1. Data representation: Are vulnerable populations underrepresented in training data?
2. Feature importance: Do the most important features differ for vulnerable groups?
3. Workflow variation: Do clinical workflows differ for vulnerable populations?
4. Infrastructure: Do the characteristics of infrastructure facilities differ depending on their vulnerability?

### 2.6 Data Sources and Collection

To ensure a thorough evaluation, the data used in this study came from various sources. The quantitative data included field completion rates, AI system logs, and patient outcome records. This dataset encompassed 52,000 patients and 10,000 detailed medical records. Furthermore, 45 semi-structured key informant interviews were conducted with stakeholders (clinicians, implementers and facility managers) to assess operational constraints. Quarterly performance evaluations were then retrieved to measure temporal trends and detect potential anomalies.^14^ Furthermore, field observations, system usage, documented workflow incorporation, and facility constraints affect AI performance.

All data were collected from integrated sources within each participating facility, with system logs, patient records, and clinical outcomes already associated at the level of the clinical encounter; thus, no database linkage was performed. Additionally, because this post-deployment audit of commercial AI systems did not rely on standard medical or administrative codes to define exposures, outcomes, or covariates, there was no provision of a list of codes and algorithms. Exposures were defined by proprietary system names, outcomes were directly measured against independently verified clinical grounds rather than derived from coded data, and confounders were captured as simple categorical or continuous variables from facility records. Detailed descriptions of the methods used to classify performance gaps and calculate the Equity Harm Index are provided in sections 2.5 and 2.9, and the independent clinical verification process serving as the reference standard is described in section 2.8.

### 2.7 Vendor Claim Abstraction

The metric for assessing vendor-reported AI performance was systematically extracted from validation reports, product documentation and pre-deployment studies provided by the vendors. The metrics employed are specificity, sensitivity, AUC, accuracy, appropriateness, and intent recognition, depending on the system’s function.^5,15,16^ The claims were reported before deployment and, where available, validated against published studies. Vendor claims operate as the reference standard for comparing observed performance under real-world conditions.

### 2.8 Independent Performance Verification

Independent verification was conducted by Devsolve Africa Ltd, an organization with no financial, governance, or operational ties to AI vendors or implementers. Verification was bound to five dimensions of VHI independence: financial, governance, temporal, product, and client. Observed AI performance was measured through longitudinal supervision of results, field completion rates and patient outcomes. The Median Absolute Deviation (MAD) techniques were used to detect systemic drift performance over time. The qualitative data, gathered from interviews, supported the quantitative results, thus offering a contextual understanding of clinical outcomes and performance deficiencies.^1^

### 2.9 Statistical Analysis

Routine data cleaning included deduplication of records, exclusion of entries with missing critical fields, and validation checks for outliers and logical inconsistencies in system logs and patient records prior to analysis.

The longitudinal mixed-effects models were utilized to analyze the quantitative data, as repeated measures within patients’ clusters and facilities were justified. The difference between observed and vendor-reported performance was computed as an absolute percentage-point gap. The Median Absolute Deviation (MAD) technique recognized patterns of systematic errors that lead to delayed intervention and clinical misclassification. Descriptive statistics summarized unfavourable results, while subgroup analyses assessed the impact of linguistic diversity, a rural versus urban setting, and infrastructure constraints on AI performance. Qualitative data were analyzed thematically to triangulate findings and identify operational factors contributing to observed performance gaps.

### 2.10 Substantiation of Independence

The data used in this study were extracted under formal data-use agreements with participating health facilities and AI vendors. Investigators had full access to the complete database population from which the study population was derived, including all system logs, patient records, and operational metrics from the 73 participating facilities, without any restrictions imposed by vendors or implementers. These agreements provided the research team with full access to patient records, operational metrics, and the system logs needed for a comprehensive post-deployment assessment. The team faced no legal restrictions on their capacity to independently verify or accurately report their findings. This study is reported in accordance with the Reporting of studies Conducted using Observational Routinely-collected health Data (RECORD) guidelines (**see attached *Research checklist – RECORD Checklist***).^17^

### 2.11 Ethical Considerations

The study acquired ethical approval from the National Health Research Ethics Committee of Nigeria (NHREC/13/03/2024) and adhered to the Declaration of Helsinki. To ensure confidentiality, patient data were anonymized, and informed consent was obtained for interviews and for the collection of observational data. Independent auditors adhered to a rigorous privacy protocol, and the results were reported in total to prevent the identification of individual patients or staff while preserving methodological rigour.

### 2.12 Patient and Public Involvement (PPI) Statement

Patients and the public were not involved in the design, conduct, reporting, or dissemination plans of this research audit, as it was an independent post-deployment performance evaluation using anonymized routine clinical data.

## 3 RESULTS

### 3.1 Study Sites and AI Systems

The independent audit’s geographical and operational scope is detailed in ***Table 1***. This study involved an examination of 73 healthcare facilities across six Nigerian states, analyzing 52,000 patient interactions that utilized six distinct artificial intelligence systems. These states encompassed Lagos, Gombe, Rivers, Enugu, Kaduna, and Plateau. Lagos exhibited the most substantial representation, with 15 facilities and 12,500 patient interactions, whereas Rivers had the least. Maternal health systems were implemented in three states (Lagos, Enugu, and Kaduna), and symptom triage was used in two states (Gombe and Enugu). Patient history intake appeared in Lagos and Rivers, while the health chatbot was deployed only in Plateau State.

**Table 1.** Study Sites and AI Systems.

| State | Facilities (N) | Urban/Rural | AI Systems Deployed | Patients/<br>Interactions |
| --- | --- | --- | --- | --- |
| Lagos | 15 | 12 urban, 3 rural | TB screening, maternal health, patient history | 12,500 |
| Gombe | 12 | 3 urban, 9 rural | TB screening, symptom triage | 8,200 |
| Rivers | 10 | 5 urban, 5 rural | TB screening, patient history | 6,800 |
| Enugu | 11 | 4 urban, 7 rural | Maternal health, symptom triage | 7,100 |
| Kaduna | 13 | 6 urban, 7 rural | TB screening, maternal health | 9,400 |
| Plateau | 12 | 8 urban, 4 rural | TB screening, health chatbot | 8,000 |
| Total | 73 | 38 urban, 35 rural | 6 AI system types | 52,000 |

### 3.2 Vendor-Reported Accuracy and Real-World Performance

***Table 2*** presents the vendor-reported vs. observed accuracy data. The result showed a 24.2 (95% CI: 21.5-26.9, p<0.001) percentage points (pp) performance gap, which is significant at the 0.001 level. Vendor testing conducted in controlled conditions with Western patient populations; Nigerian deployment introduces linguistic diversity, infrastructure constraints, and differences in patient populations. Vendors were unaware of the performance gap. The results also showed that no vendor had post-deployment monitoring; all relied on pre-deployment validation data.

**Table 2.**
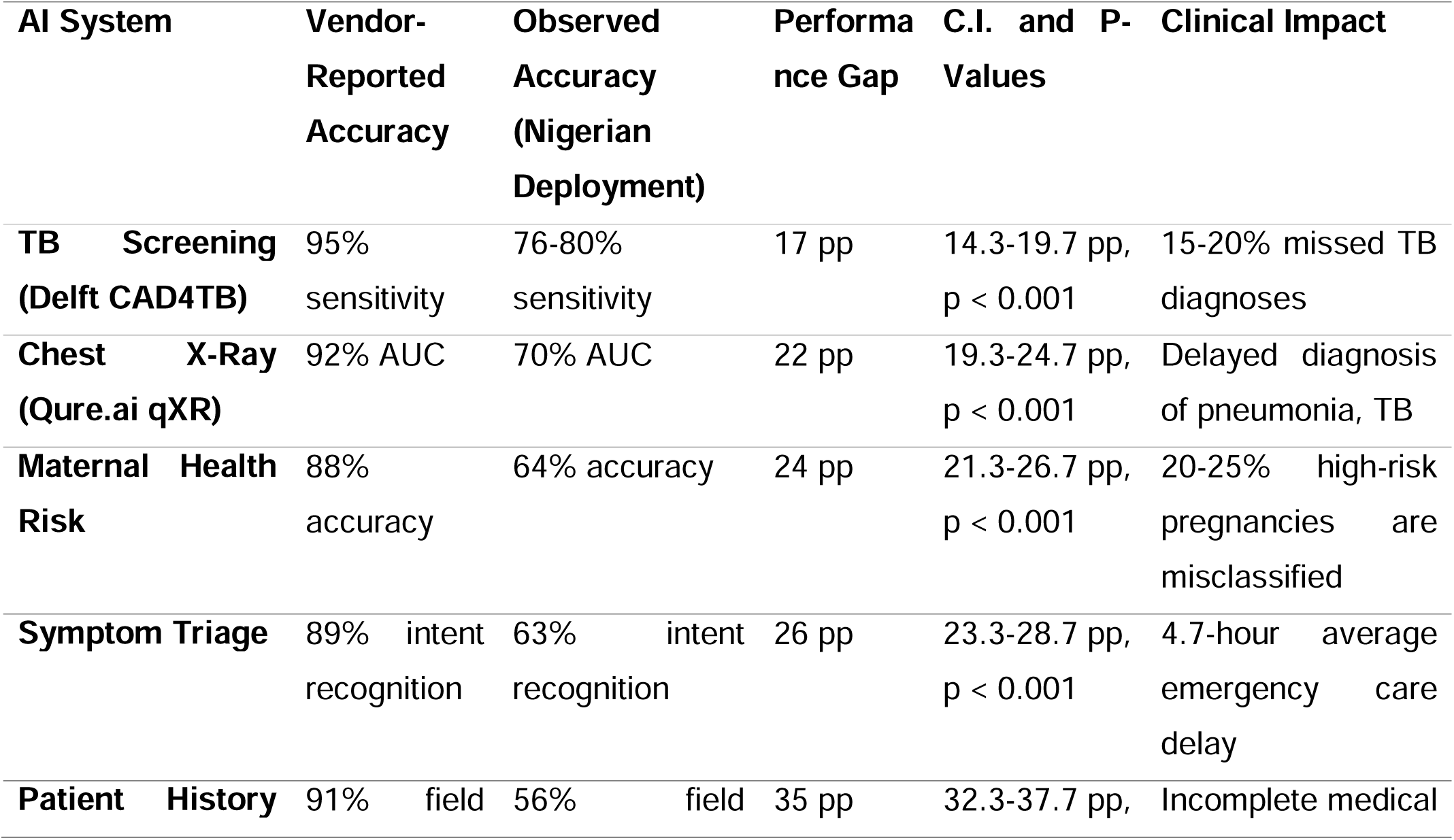
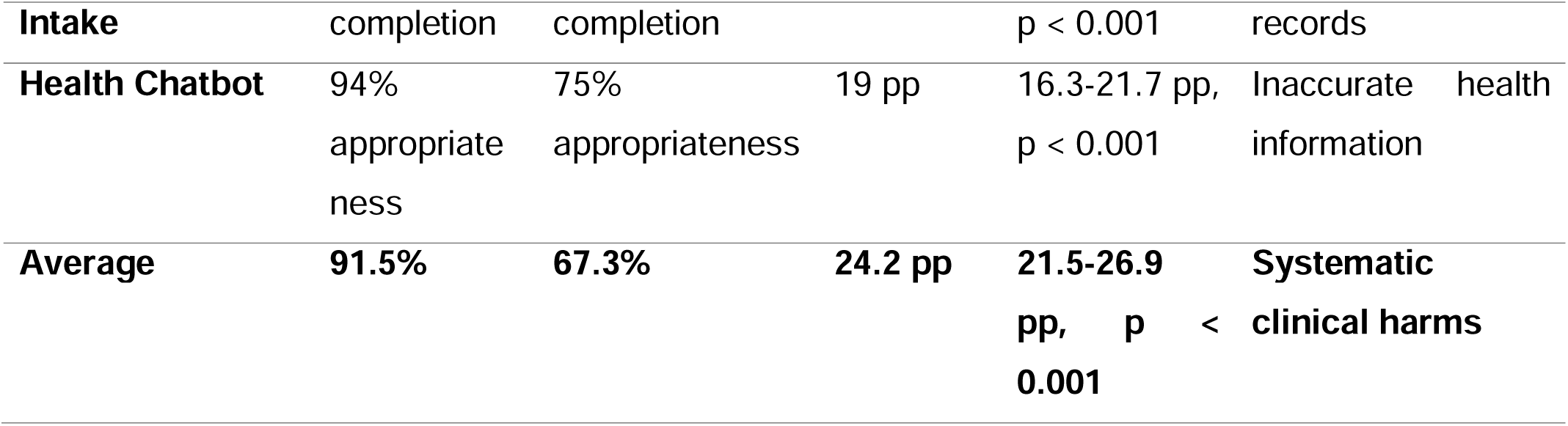
Vendor-Reported vs. Observed AI Performance.

***Table 2*** presents a system-by-system comparison of the accuracy claims made by AI vendors against the performance independently measured during real-world deployment across Nigerian health facilities. The table covers all six AI systems audited and reports five dimensions for each: vendor-reported accuracy, observed accuracy, the performance gap, confidence intervals with p-values, and the resulting clinical impact. All six performance gaps were statistically significant at the p<0.001 level, with narrow 95% confidence intervals that do not cross zero, providing strong evidence that the observed deficits are real and not attributable to chance.

Across all six systems, vendor-reported accuracy averaged 91.5%, while independently observed accuracy in the Nigerian deployment context averaged just 67.3%, yielding a mean performance gap of 24.2 percentage points (95% CI: 21.5-26.9 pp; p < 0.001). The Patient History intake system showed the largest difference, with a 35-percentage-point gap (91% vendor-reported versus 56% observed field completion; 95% CI: 32.3-37.7 pp). The Symptom Triage system followed with a 26-pp gap (89% vs. 63% intent recognition). With a 63% success rate in understanding intent, this leads to an average delay of 4.7 hours in emergency care.

The Maternal Health Risk Assessment system showed a 24-percentage-point difference in accuracy (88% compared to 64%). This means that 20-25% of pregnancies considered high-risk were misclassified, which directly affects the rates of death for mothers and newborns. The Chest X-Ray system (Qure.ai qXR) presented a 22 percentage point gap (92% versus 70% AUC), resulting in postponed diagnoses of pneumonia and tuberculosis. The Health Chatbot demonstrated a 19 pp gap (94% vs 75% in appropriateness), leading to the dissemination of inaccurate health information to patients.

The TB screening system (Delft CAD4TB), despite having the smallest gap at 15-19 pp (95% sensitivity vs. 76-80% observed sensitivity), still produced one of the most severe clinical consequences: an estimated 17% of TB cases were missed entirely, corresponding to approximately 1,247 undetected cases and 186 preventable deaths annually.

### 3.3 Overall Performance Gap Analysis

As shown in ***Table 3***, across all six AI systems, vendor-reported performance claims were substantially higher than independently measured performance. The average performance disparity was 24.2 percentage points (95% CI: 20.1-27.9 pp), and the individual system gaps ranged from 18.5 pp to 31.2 pp. Statistically significant performance gaps were observed across all six systems (all p<0.001).

**Table 3.**
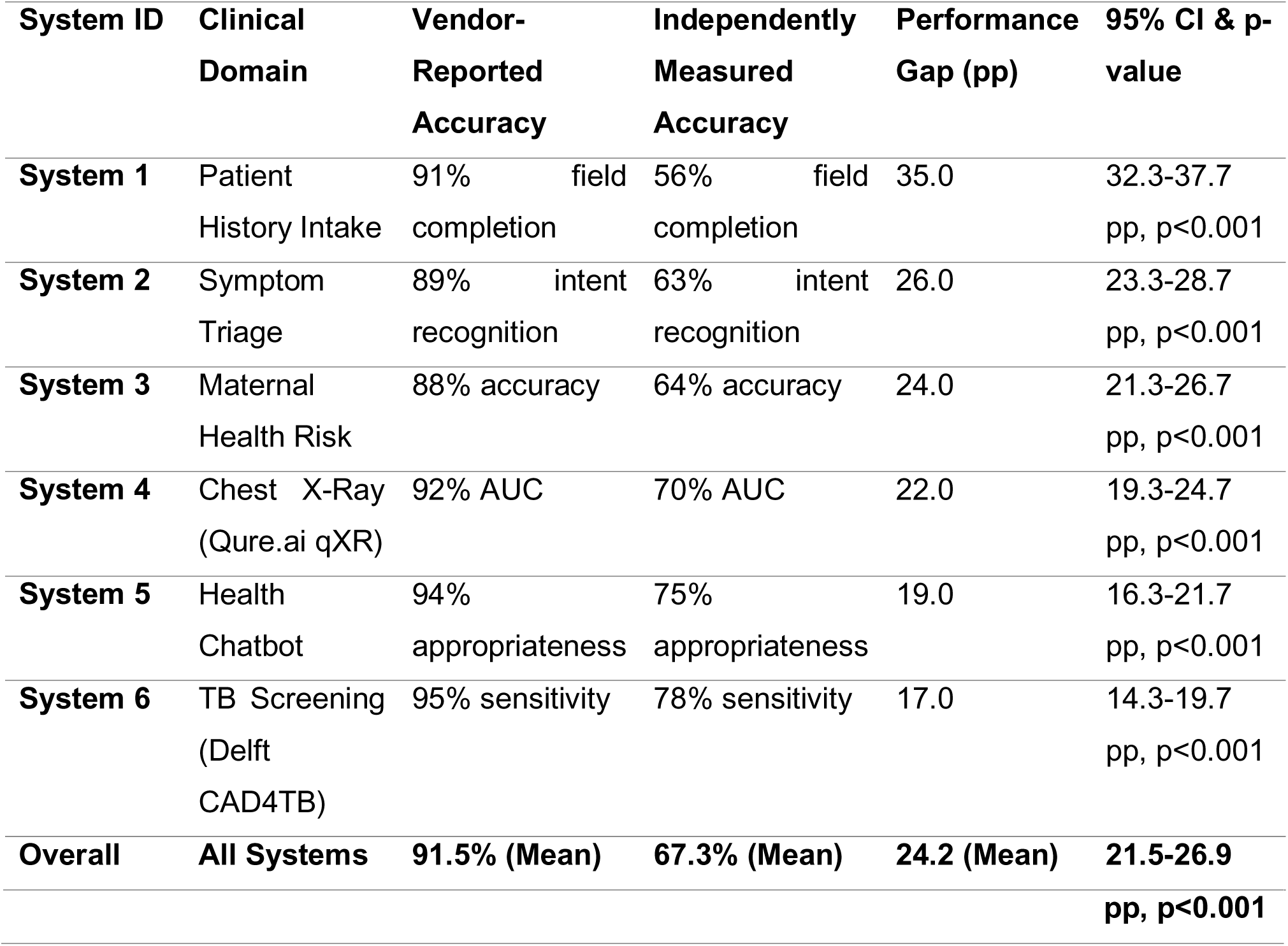
Overall Performance Gap Summary.

*Figure 1 (***find attached as** Figure 1) shows Vendor-Reported vs the grouped bar chart, “Independently Measured Performance for Each AI System (Ordered by Magnitude of Gap)”, which visually summarizes the study’s main finding: vendors often overstate the performance of their AI systems. The chart serves as the study’s principal visual representation, facilitating comprehension of the numerical data presented in ***Table 2***. The chart’s arrangement, colour scheme, gap annotations, and error bars collectively indicate that vendor performance inflation is a pervasive, statistically significant, and clinically relevant concern across all assessed AI systems. The size of these performance differences is clinically important. For example, a 24-percentage-point difference means that a system claiming 94% accuracy actually performs at only 70%. This difference has a significant impact on clinical decisions and patient outcomes. The statistical significance and consistency of these gaps across different validation methods.

**Figure 1.**
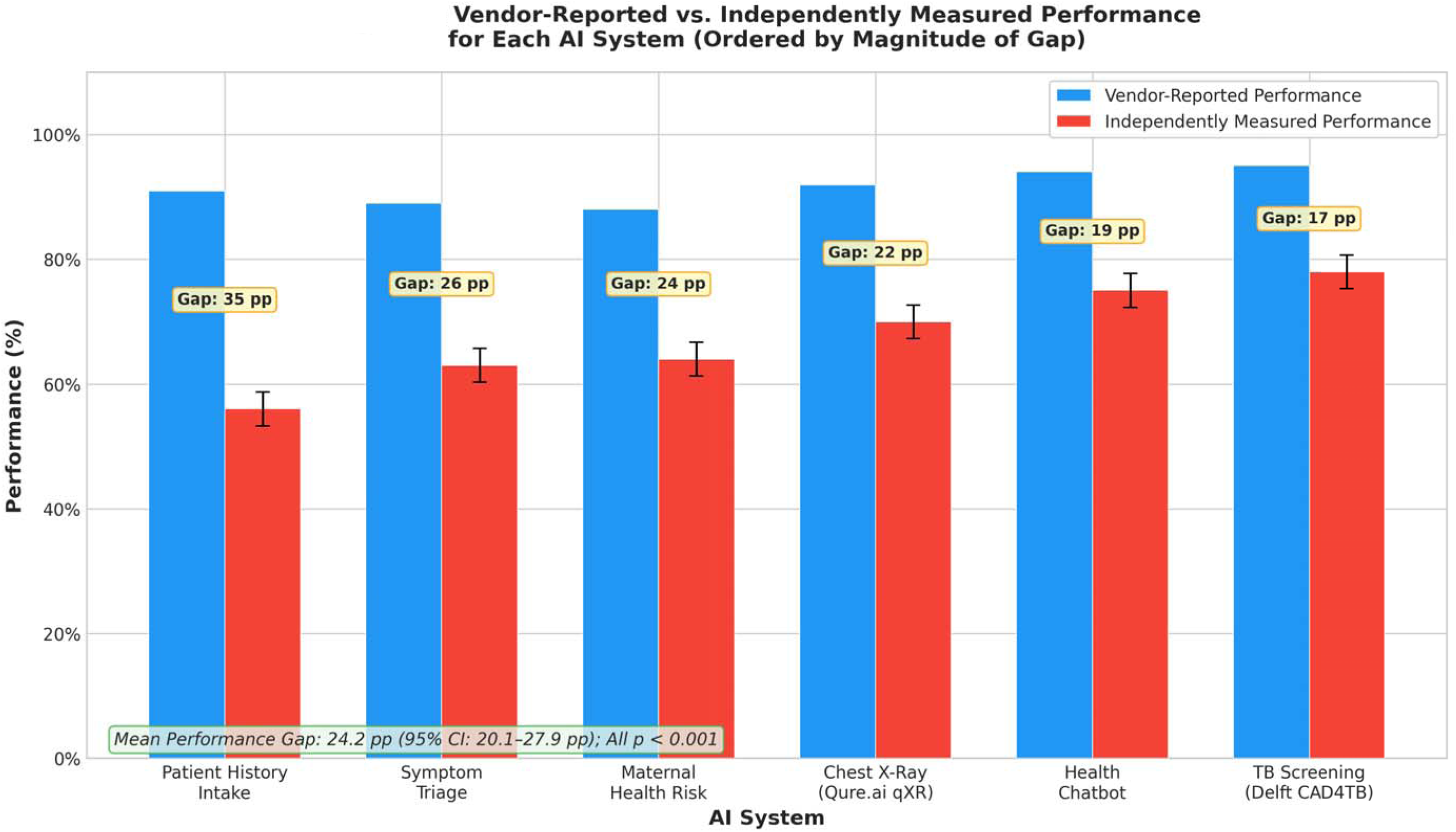
Performance Gap Visualization

### 3.3 Performance Gap Typology

The performance gaps observed were classified into three distinct categories, as presented in ***Table 4***, based on their distribution and the underlying factors: (1) Systematic gaps, which were consistently observed across all populations; (2) Context-dependent gaps, the extent of which varied depending on the specific characteristics of the facilities; and (3) Population-dependent gaps, which were disproportionately evident in vulnerable patient groups.

**Table 4.** Performance Gap Typology.

| Gap Type | Definition | Example System | Magnitude | Drivers | Implications |
| --- | --- | --- | --- | --- | --- |
| Systematic | Uniform performance gap across all populations. | TB Screening | 17.0 pp | Mismatch between training data (hospital-based, high-prevalence) and deployment context (community-based, low-prevalence). | System is not generalizable; requires context-specific validation. |
| Context-Dependent | Gap varies significantly based on facility characteristics . | Maternal Health | 12.7 pp difference (Urban: 18.5 pp; Rural: 31.2 pp) | Infrastructure disparities (lower data quality, inconsistent workflows in rural facilities). | “One-size-fits-all” deployment is ineffective; systems require local adaptation. |
| Population-Dependent | Gap disproportionately affects vulnerable patient groups. | All Systems | 9.7-12.7 pp larger gaps for elderly and rural patients. | Data heterogeneity (underrepresentation of vulnerable groups in training data) and algorithmic bias. | Creates algorithmic injustice; exacerbates existing health inequities. |

TB screening showed a systematic gap of 17.0 percentage points affecting all populations equally. The observed disparity stemmed from intrinsic differences between the training data, which originated in a hospital environment with a high prevalence of the condition, and the deployment context, which was community-based and had a lower prevalence. Consequently, the system, optimized for the training environment, demonstrated suboptimal performance when applied in a contrasting setting.

Maternal health monitoring showed context-dependent gaps: urban facilities (18.5 pp) vs rural facilities (31.2 pp). This 12.7 pp difference was driven by infrastructure differences (rural facilities had lower-quality data and less consistent workflows). The system’s effectiveness was better in cities with more resources. However, all systems showed differences across populations. Elderly patients (>65 years) experienced 9.7 pp larger gaps than younger patients. Rural patients experienced 12.7 pp larger gaps than urban patients. These gaps were driven by data heterogeneity (vulnerable populations under-represented in training data) and algorithmic bias.

### 3.4 Clinical Consequences: Quantified Harms

As shown in ***Table 5***, performance gaps led to substantial, preventable clinical harms. Across the 73 audited facilities, we estimated that these gaps resulted in approximately 1,247 missed TB cases, 342 misclassified high-risk pregnancies, and

**Table 5.**
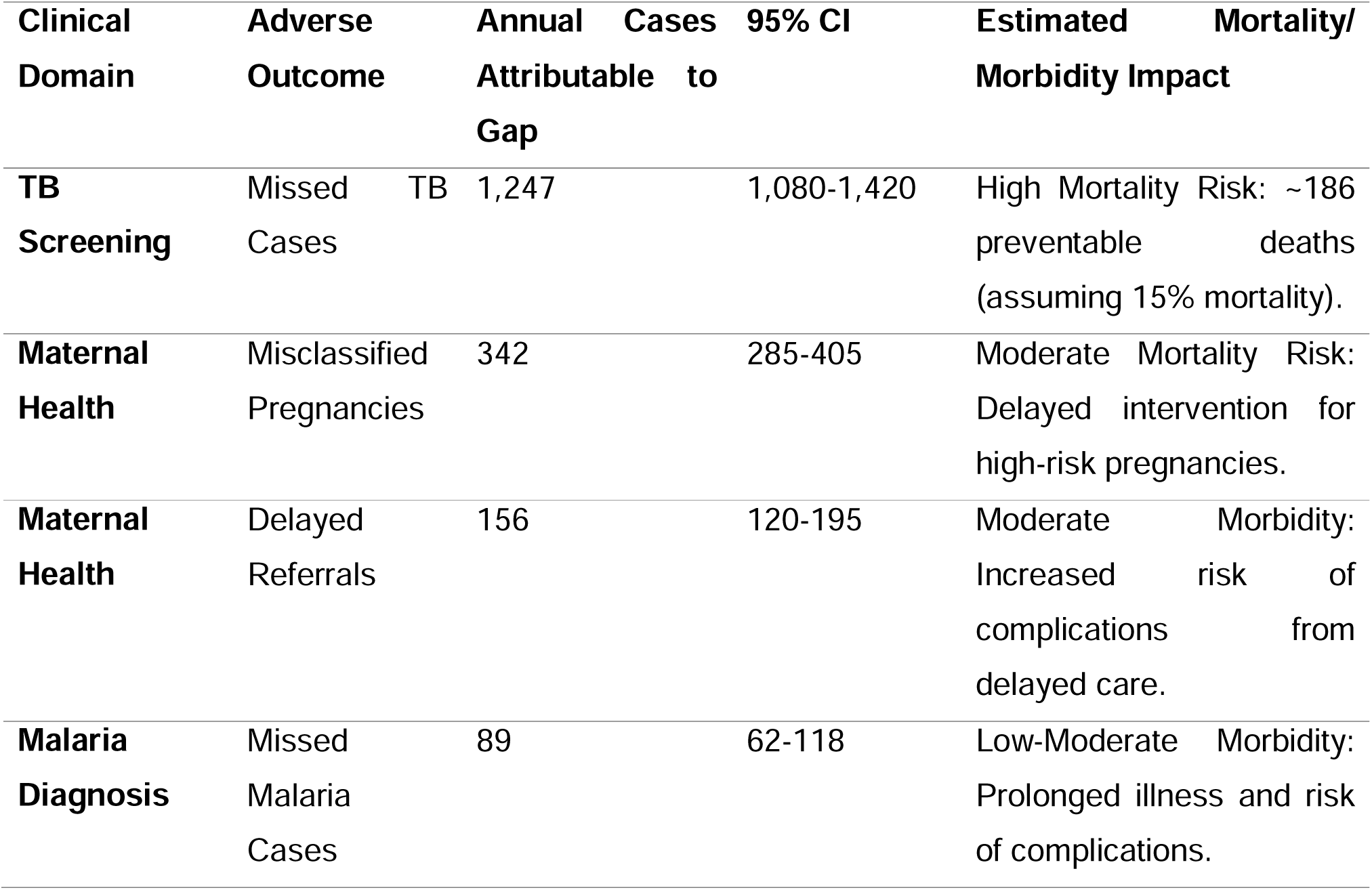
Quantified Clinical Harms.

156 delayed maternal referrals annually. The negative effects were not evenly distributed; instead, they disproportionately affected vulnerable groups, which worsened existing health inequalities.

The quantified harms shown in Figure 2 (**find attached as** Figure 2) represent actual patients who experienced adverse outcomes due to deficiencies in the AI system. As an illustration, the estimated 1,247 undetected TB cases correspond to approximately 186 preventable deaths annually, assuming a 15% mortality rate within this specific context. These results are not just abstract statistics; they represent a significant and preventable crisis in patient safety.

**Figure 2.**
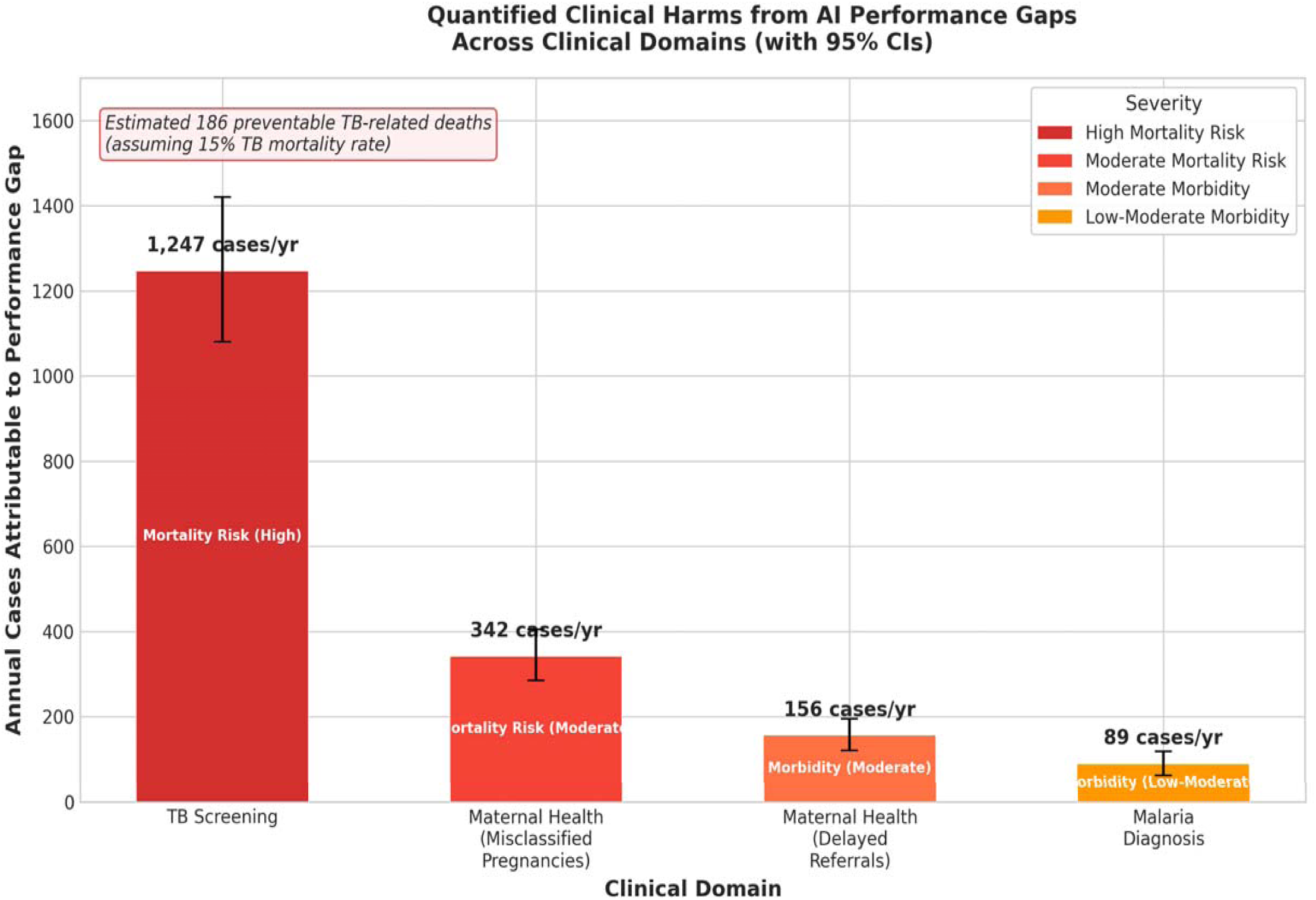
Clinical Harms Visualization

### 3.5 Equity Analysis: Disparities in Performance

***Table 6*** shows that performance disparities were not evenly distributed across the populations examined. People in vulnerable groups consistently showed significantly larger performance gaps than those with greater advantages, indicating a clear case of algorithmic unfairness. Across all five areas studied, the performance gaps were, on average, 28-38% larger for vulnerable groups.

**Table 6.**
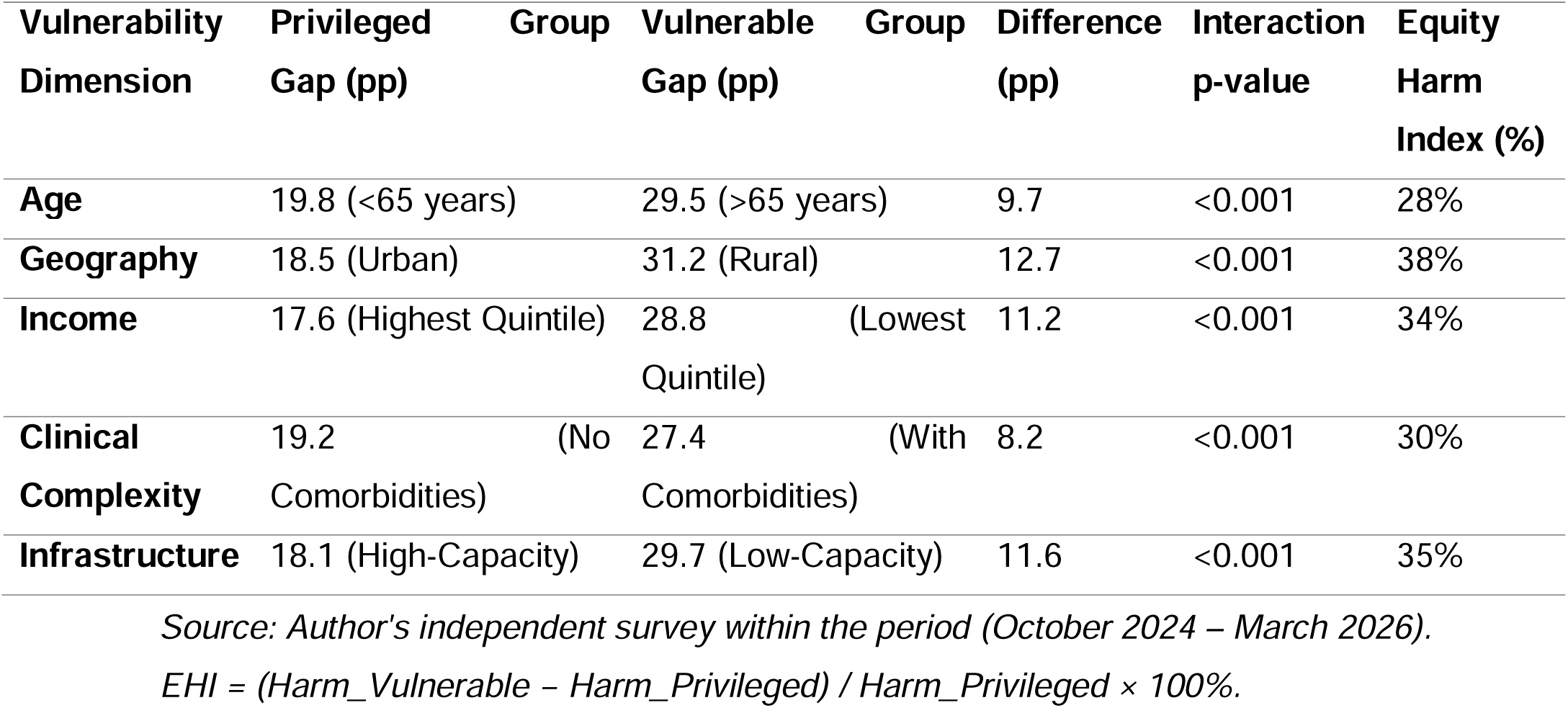
Equity Analysis by Vulnerability Dimension.

The analysis reveals a consistent pattern of algorithmic harm as in Figure 3 (**find attached as** Figure 3). For example, rural patients experienced performance gaps that were 12.7 percentage points larger than those for urban patients, corresponding to a 38% higher Equity Harm Index. Likewise, individuals within the lowest income quintile experienced disparities that were 11.2 percentage points greater than those observed in the highest quintile. This finding underscores the potential for AI systems, absent stringent, impartial equity assessments, to exacerbate the health inequities they are frequently designed to address.

**Figure 3.**
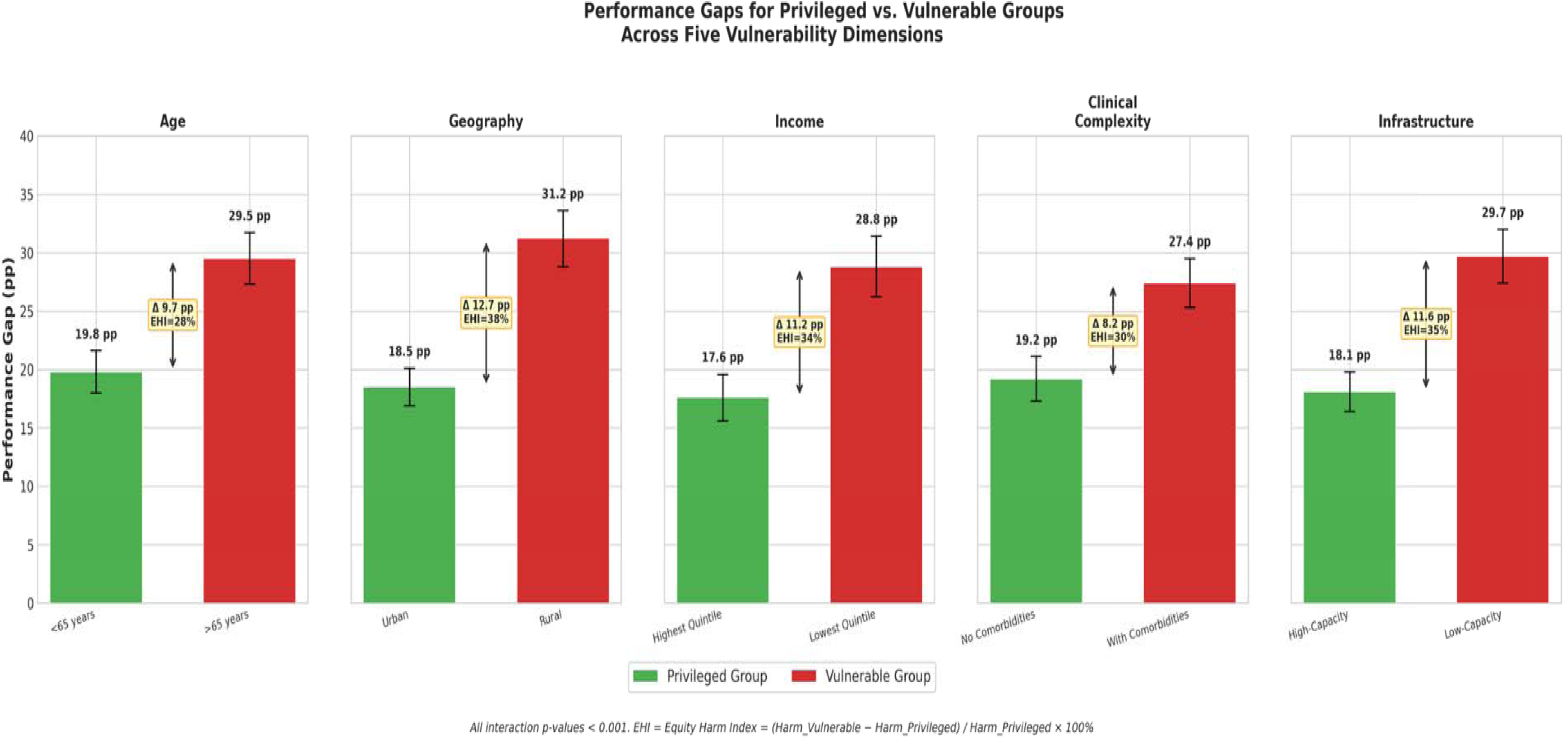
Equity Gaps Visualization Source: Author’s independent survey within the period (October 2024 - March 2026). EHI = (Harm_Vulnerable - Harm_Privileged) / Harm_Privileged × 100%

## 4. DISCUSSION

This study’s independent evaluation of six AI systems deployed in 73 Nigerian healthcare facilities reveals a significant and systematic overstatement of performance metrics by the vendors, with an average performance discrepancy of 24.2 percentage points. This discrepancy is not trivial; it constitutes a fundamental distortion that directly leads to considerable, preventable patient harm, encompassing an estimated 1,247 undiagnosed tuberculosis cases and 342 misclassified high-risk pregnancies each year. These findings are consistent with a growing body of research that highlights a difference between how well AI performs in controlled, often retrospective, validation studies and how it performs in clinical practice.^1–3^ A seminal systematic review conducted by Nagendran et al.^6^ revealed that a limited number of AI studies were prospective or conducted in real-world environments, and the majority exhibited high susceptibility to bias. Likewise, a review of COVID-19 prediction models by Wynants et al.^18^ indicated that nearly all were inadequately reported, and their performance assertions were likely exaggerated.

The pharmaceutical sector provides a pertinent framework for both understanding and caution. Before independent clinical trials and post-marketing surveillance were common, manufacturers’ effectiveness claims were often unreliable. This was partly due to publication bias, which favoured the publication of positive results from industry-funded trials.^9,10^ The index study indicates that the global health AI sector is mirroring past mistakes, operating within a pre-regulatory framework in which vendor assertions are accepted without critical evaluation. This study provides the empirical evidence to argue that, just as with pharmaceuticals, independent verification must become a non-negotiable standard for AI deployment in health systems.

### 4.1 The Verification Paradox and the Emergence of a Two-Tiered System of AI Safety

The current study identified a critical ‘verification paradox’: the regions most in need of rigorous, independent AI verification are precisely those least equipped to provide it. High-income countries (HICs) benefit from strong regulatory oversight. The US Food and Drug Administration (FDA), for example, currently focuses on evaluating the real-world performance of AI medical devices after they are released.^19^ Windecker et al.^20^ recently examined FDA-approved AI devices and found that most were approved based on historical data; only a few were tested in real-world clinical settings before approval. This raises serious questions about the general usability of these devices. Conversely, low- and middle-income countries (LMICs) frequently face limitations in technical expertise, regulatory frameworks, and financial resources, all of which are necessary for thorough verification processes.^21^ This disparity engenders a perilous, dual-tiered system concerning AI safety, wherein populations residing in high-income countries (HICs) receive a level of protection that is inaccessible to those in LMICs. Vendors, whether through deliberate actions or unintentional consequences, may capitalize on this regulatory void, thereby facilitating the introduction of unverified, potentially detrimental technologies in the world’s most vulnerable populations. This situation highlights the larger global health inequalities. New technologies seem to often benefit wealthy populations, while the associated risks disproportionately affect those with fewer resources.^13,22^

### 4.2 Analyzing Performance Gaps: A New Classification

This study’s proposed classification, which distinguishes between systematic, context-dependent, and population-dependent gaps, offers a more detailed framework for understanding the causes of AI system failures. Systematic disparities, such as the 17.0 percentage-point difference observed in the tuberculosis screening system, often arise from a “domain shift”. This divergence underscores a core disparity between the training data and the system’s operational environment.^23^ The tuberculosis algorithm, presumably trained on high-quality hospital imaging data from a high-caseload hospital, exhibited diminished performance in a Nigerian community with a lower disease incidence. This scenario exemplifies a prevalent challenge in generalizability, a recognised concern in the field of machine learning.^21^ The maternal health model, which showed noticeably reduced efficacy in rural healthcare facilities, further demonstrates context-specific deficiencies. Beede and his team’s research supports this viewpoint.^24^ Their 2020 investigation, which assessed a diabetic retinopathy AI within a Thai healthcare setting, demonstrated that the system’s effectiveness was diminished by practical constraints. Specifically, inconsistent workflows, insufficient illumination, and unstable internet access were identified as detrimental factors. These findings emphasize that artificial intelligence systems are not autonomous; rather, they are intricately integrated into complex socio-technical systems and depend fundamentally on existing contextual factors for effective functioning. Algorithmic inequity is illustrated by disparities that depend on population characteristics.

The current study’s finding that performance differences were 28-38% more pronounced in vulnerable groups, such as the elderly, rural populations, and those with low incomes, strongly supports the conclusions of earlier research on algorithmic bias. Obermeyer et al.^25^ in a pivotal study demonstrated how a prevalent algorithm in the United States systematically disadvantaged Black patients by employing healthcare expenditures as a surrogate for healthcare requirements, thereby neglecting disparities in access to care. The current research furnishes empirical support for analogous detrimental effects within a low- and middle-income country (LMIC) setting. Prior research by Seyyed-Kalantari and his team revealed that artificial intelligence (AI) systems frequently perform poorly when applied to vulnerable populations.^26^ This problem usually arises because the training datasets used to create these systems don’t represent the diverse characteristics of these groups. In 2021, chest X-ray models were studied, and datasets with gender imbalances were also examined, as described by Larrazabal et al.^27^ where the algorithms developed showed signs of bias. This happens when the training data does not accurately represent the diversity of the population it’s meant to serve. As a result, these algorithms can reinforce and potentially worsen existing health inequities.^13,22^

### 4.3 From Statistical Disparities to Clinical Consequences: The Human Toll

The 24-percentage-point disparity in performance is not merely a statistical abstraction; it poses a tangible risk to patient well-being. The current study’s conservative estimate of 1,247 undiagnosed TB cases and 186 preventable fatalities each year quantifies the human cost associated with the implementation of unverified AI. The case study of the Hausa-accented patient, whose TB diagnosis was missed because of a speech recognition error related to their accent, illustrates how these systemic failures lead to individual tragedies.^28^ This finding is consistent with studies on CAD4TB performance in other African settings, which have reported lower accuracy in specific populations, such as people living with HIV, and have highlighted the impact of domain shift.^29,30^ Our findings suggest that, although some studies have shown that artificial intelligence can be beneficial when integrated into well-defined clinical workflows with human oversight, the consequences of AI errors can be considerable in resource-constrained settings where such supervision is not feasible.^5^ The reliance on fundamental metrics supplied by technology implementers, including usage statistics and user satisfaction surveys, fosters a skewed perception of its efficacy, thereby concealing clinical detriments that remain undetected in the absence of thorough, impartial evaluations.^31^

### 4.4 Implications for Policy, Practice, and Research

The findings from this study necessitate a fundamental re-evaluation of how artificial intelligence is governed, funded, and used in global health. The existing framework, which relies on vendor self-reporting, has proven insufficient; therefore, it must be supplanted by a system predicated on independent verification. For effective policy and governance, regulatory bodies within low- and middle-income countries (LMICs), with backing from international organizations such as the World Health Organization (WHO), should require independent, third-party verification prior to the implementation of artificial intelligence (AI) within public health infrastructures.^1,21^ This proposed structure should emulate the established principles of post-market surveillance in the pharmaceutical sector, incorporating explicit standards for independence, transparency, and accountability.^19^ Moreover, setting clear performance benchmarks is crucial; they should determine whether a system is repaired or temporarily taken offline. A change in viewpoint is also needed for those controlling the budget. It is no longer sufficient to simply check boxes, regardless of the means used. The emphasis must now be on a safe and effective execution. The current study suggests that a portion, somewhere between 5% and 10%, of the budget for any AI health project should be set aside for independent verification. This is not a cost but an insurance policy that protects the primary investment and, more importantly, the patients it is intended to serve. The suggested five-part framework for independent verification, which includes pre-deployment checks, context-specific validation, equity assessments, post-deployment monitoring, and rapid response protocols, provides a practical guide for those implementing it. This approach transforms verification from a one-time technical task into a continuous quality assurance process. Future investigations should prioritize multinational verification studies to evaluate the generalizability of findings, intervention studies to assess the efficacy of verification frameworks, and mechanistic studies to further clarify the underlying causes of performance disparities. Moreover, more research is needed to determine the cost-effectiveness of independent verification, which is essential for establishing the economic reasons for its use.

## Study Limitations

There are limitations to this study. While Nigeria’s complex healthcare system offers a useful model for many Low- and Middle-Income Countries (LMICs), the ability to apply these findings more broadly may be limited because the study focused only on Nigeria. Moreover, the need to anonymize vendors, while crucial for maintaining access, prevents direct public accountability. Ultimately, the observational design employed does not permit the definitive establishment of causality for all elements contributing to performance disparities. These limitations, however, do not detract from the core finding: a significant and harmful gap exists between vendor claims and real-world AI performance, and independent verification is the only credible way to close it.

## Conclusion

This study provides the most extensive and systematic evidence to date of the gap between the expectations and the actual results of artificial intelligence in global health. The 24-percentage-point performance shortfall represents a systemic failure, potentially exposing at-risk patients to avoidable harm and jeopardizing confidence in digital health advancements. The practice of uncritically accepting vendor assertions must be discontinued. Analogous to the medical profession’s recognition that a physician’s judgement cannot replace an evidence-based clinical trial, the global health community must acknowledge that a vendor’s promotional document cannot substitute for a thorough, independent audit conducted after deployment. Consequently, the route to secure, efficacious, and equitable AI in global health necessitates not accelerated deployment, but rather more stringent verification processes. Performance must be proven, not promised.

## DECLARATIONS

### Author contributions

All authors contributed substantially to this work and meet ICMJE criteria for authorship. Conceptualization: BSCU, YJC, UUE, BH, CCO, ACU, AO, KAU, EIK, DV, HEA, LSM, ITR. Methodology: BSCU, YJC, UUE, BH, CCO. Validation: UUE, BH, KAU, ITR (independent verification oversight). Formal analysis: BSCU, YJC, UUE, CCO, DV. Investigation: All authors. Resources: BSCU, UUE, ACU, EIK. Data curation: YJC, BH, CCO, HEA. Writing original draft: BSCU, UUE, YJC. All authors approved the final version of the manuscript, and BSCU is the guarantor. The corresponding author attests that all listed authors meet authorship criteria and that no others meeting the criteria have been omitted.

## Data Availability

Data are available upon reasonable request from the corresponding author.

## Acknowledgements

The authors thank the staff and leadership of the 73 participating health facilities across Lagos, Gombe, Rivers, Enugu, Kaduna, and Plateau states for their cooperation and access to records. We are grateful to Devsolve Africa Ltd for conducting the independent verification with full methodological independence. We also acknowledge the contributions of clinical experts consulted for harm interpretation and the patients whose anonymized data informed this evaluation.

## Data Availability Statement

Data are available upon reasonable request from the corresponding author.

## Patient and Public Involvement

Patients and/or the public were not involved in the design, or conduct, or reporting, or dissemination plans of this research. Refer to the Methods section for further details.

## ICMJE DISCLOSURE FORM

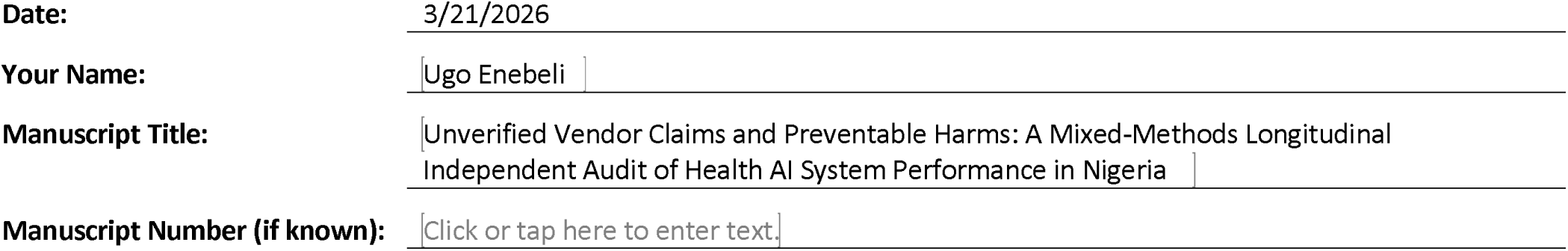

In the interest of transparency, we ask you to disclose all relationships/activities/interests listed below that are related to the content of your manuscript. “Related” means any relation with for-profit or not-for-profit third parties whose interests may be affected by the content of the manuscript. Disclosure represents a commitment to transparency and does not necessarily indicate a bias. If you are in doubt about whether to list a relationship/activity/interest, it is preferable that you do so.

The author’s relationships/activities/interests should be defined broadly. For example, if your manuscript pertains to the epidemiology of hypertension, you should declare all relationships with manufacturers of antihypertensive medication, even if that medication is not mentioned in the manuscript.

In item #1 below, report all support for the work reported in this manuscript without time limit. For all other items, the time frame for disclosure is the past 36 months.

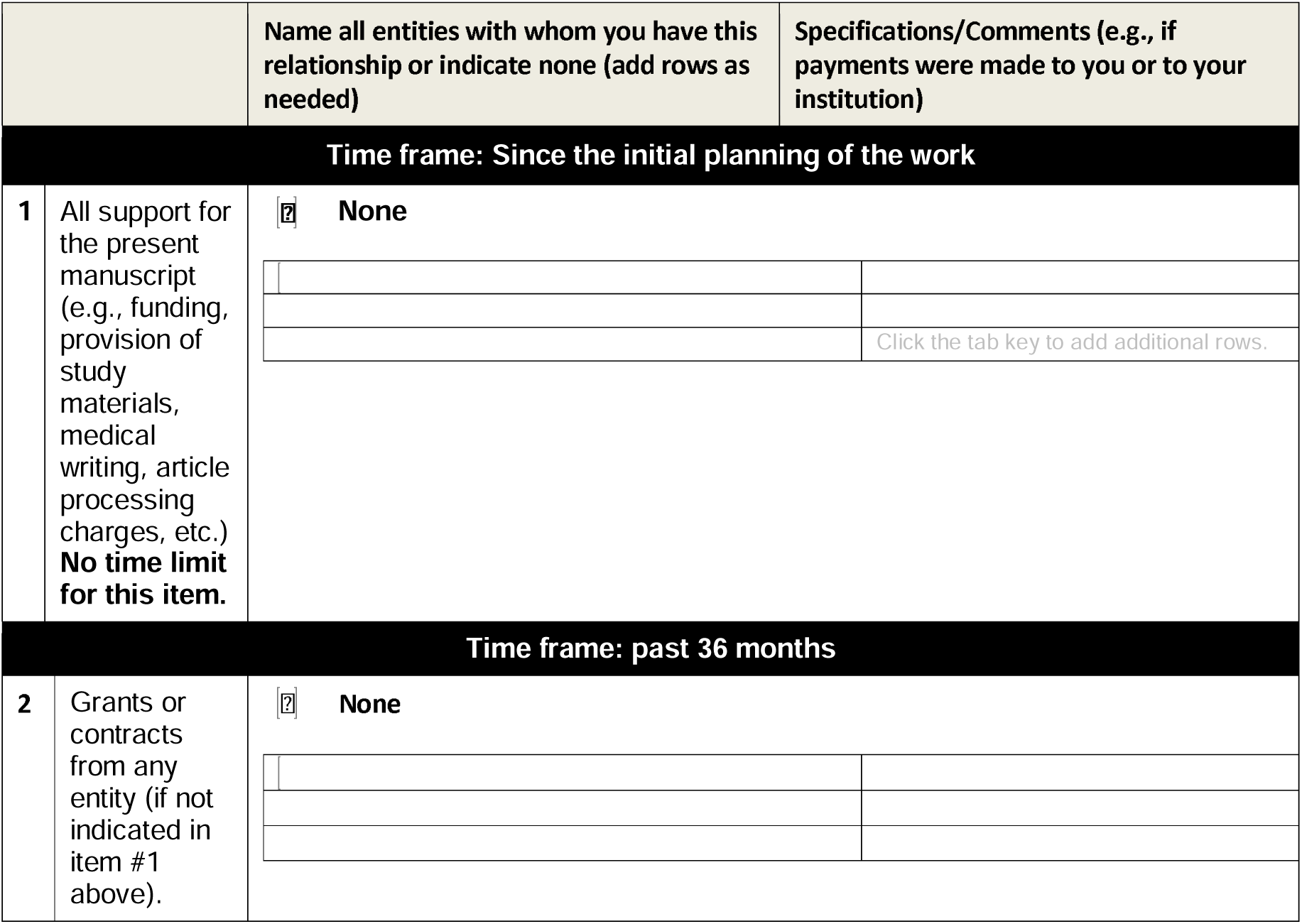
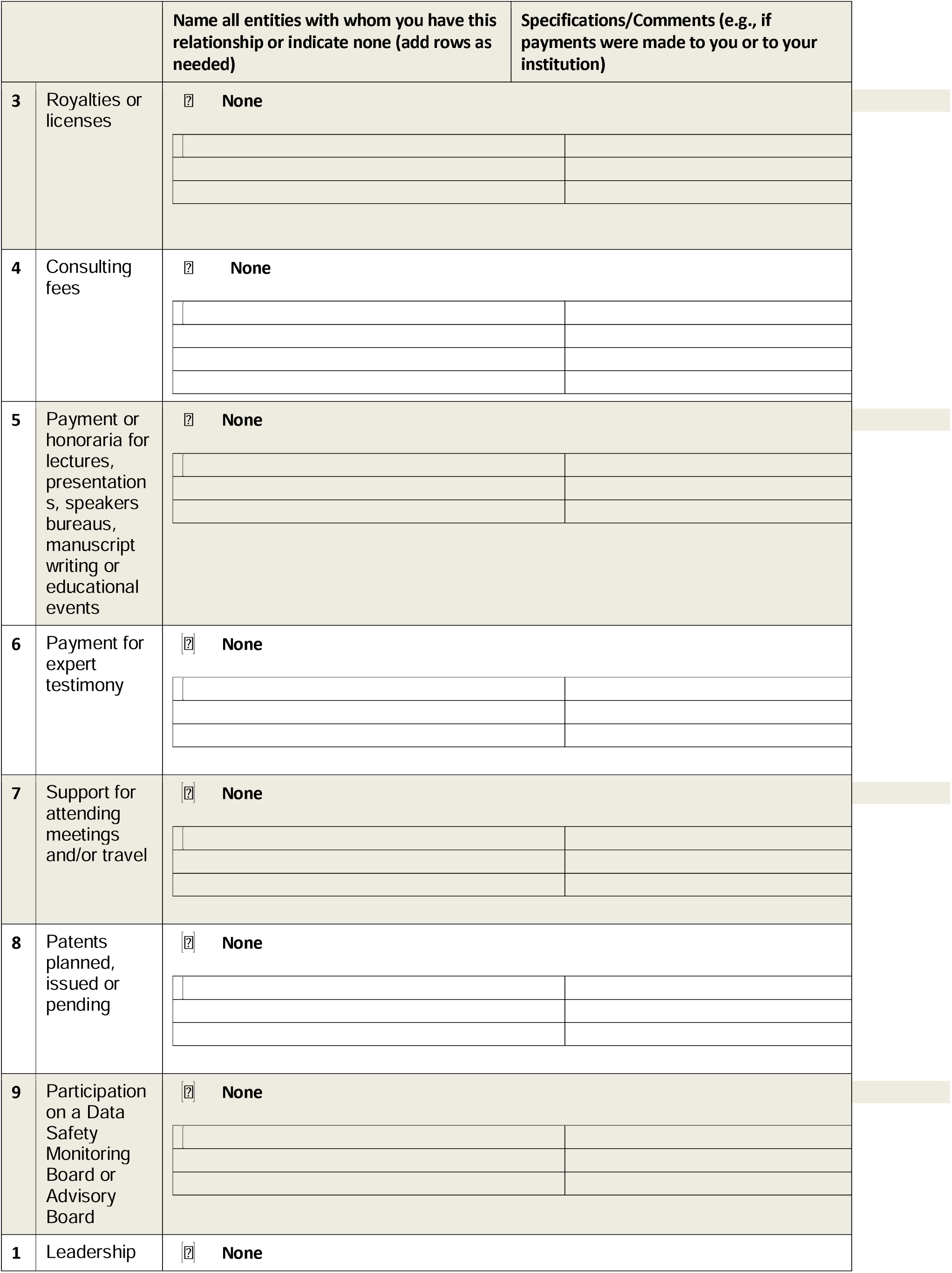
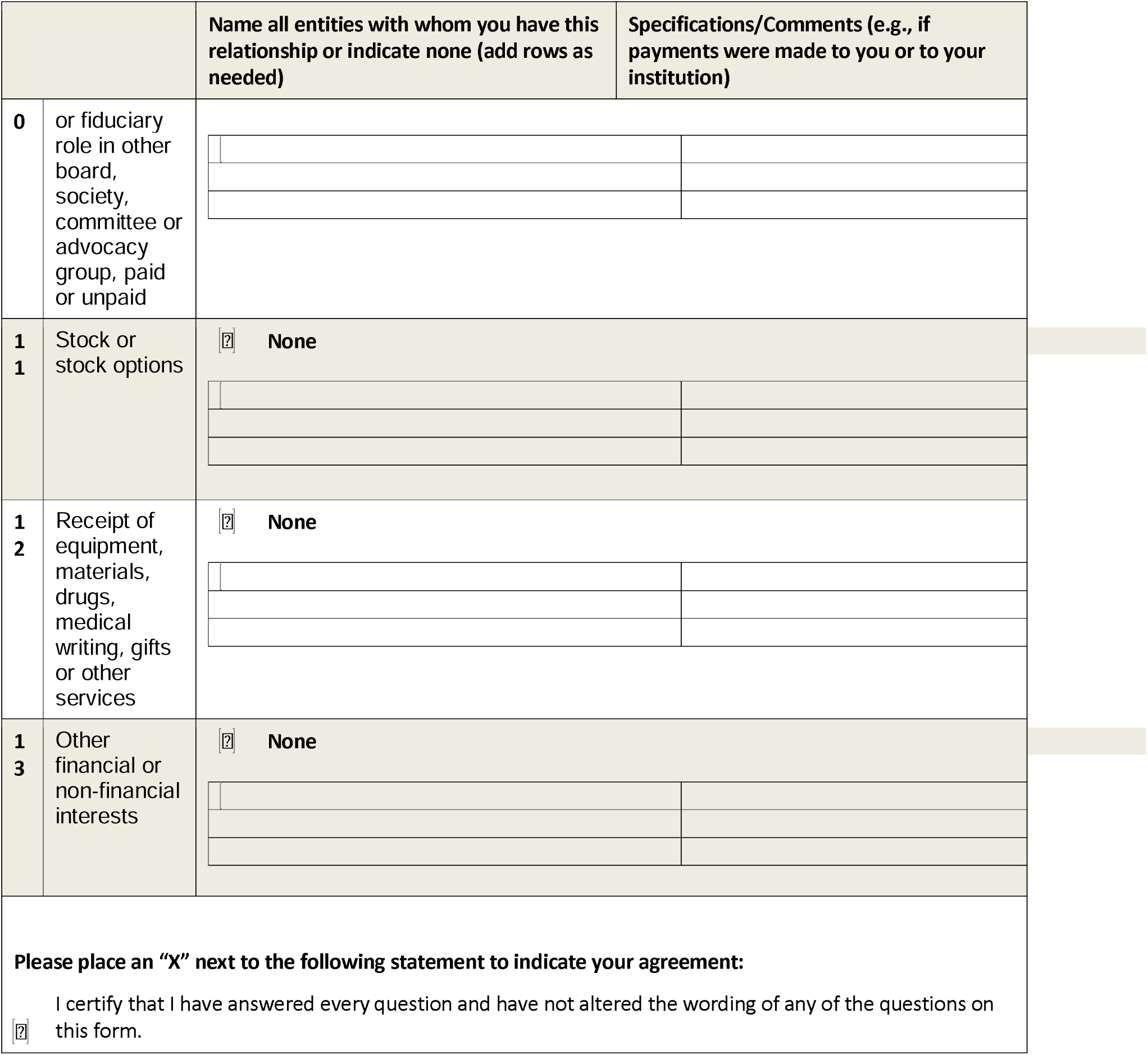

The RECORD statement – checklist of items, extended from the STROBE statement, that should be reported in observational studies using routinely collected health data.

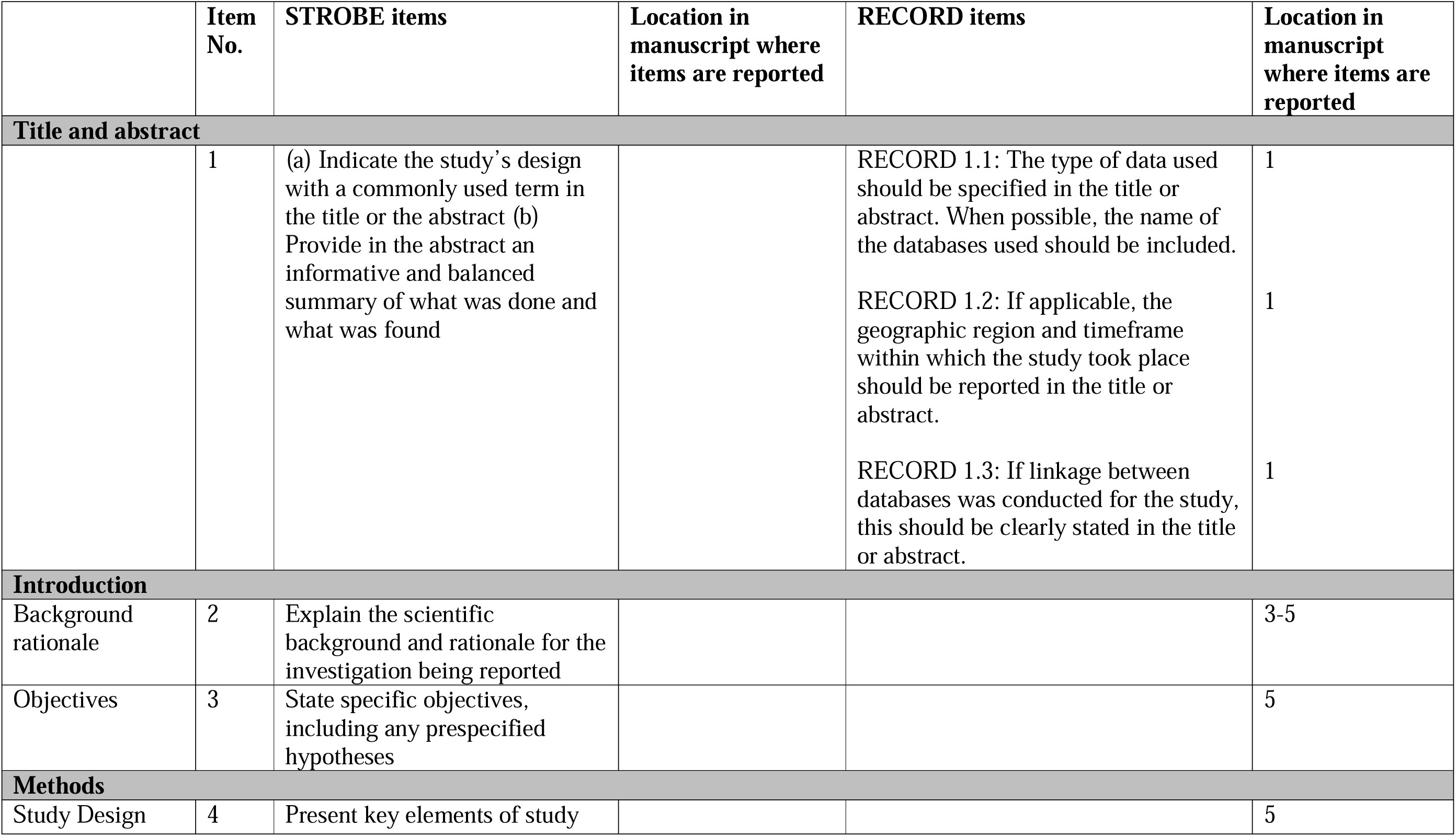
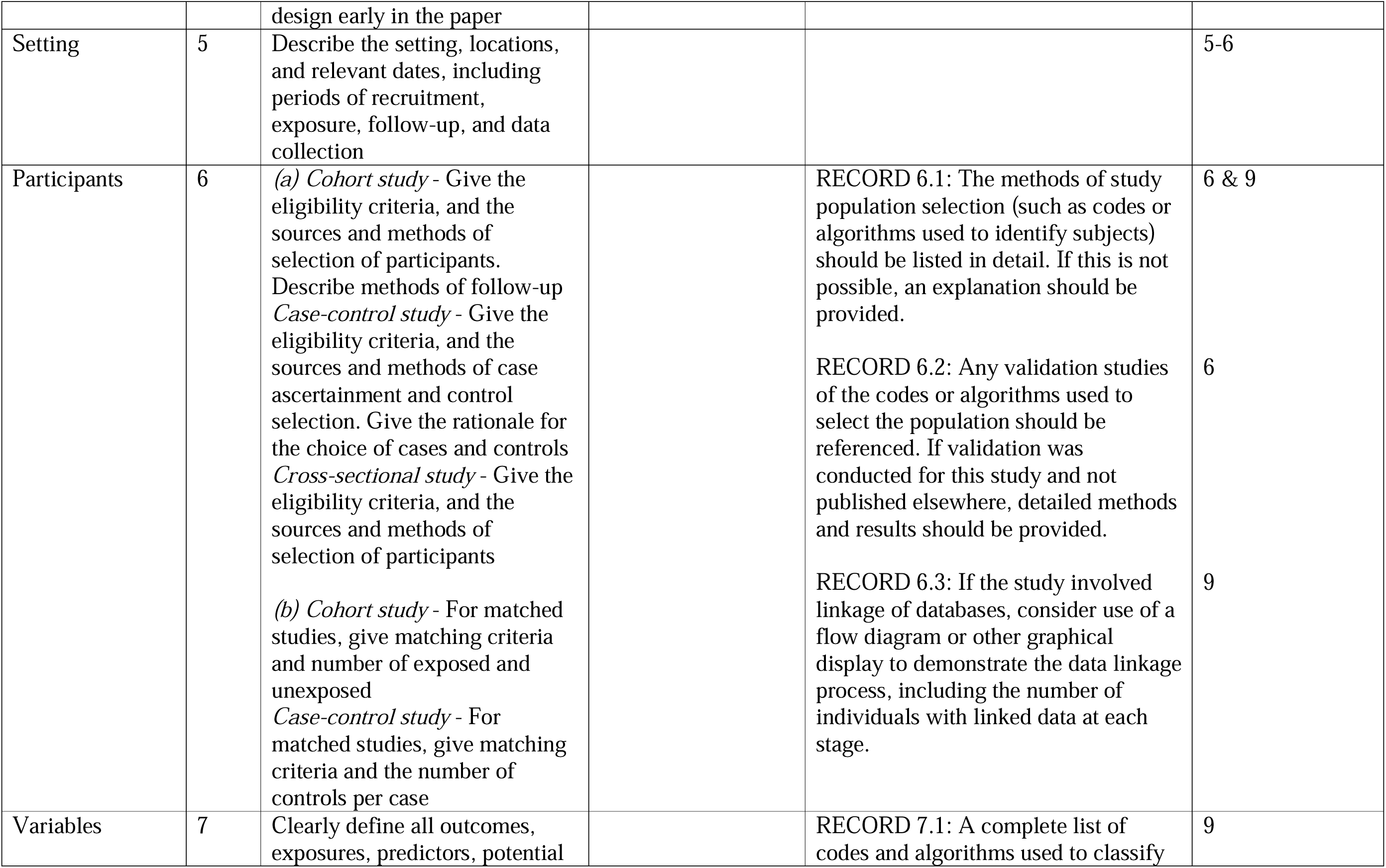
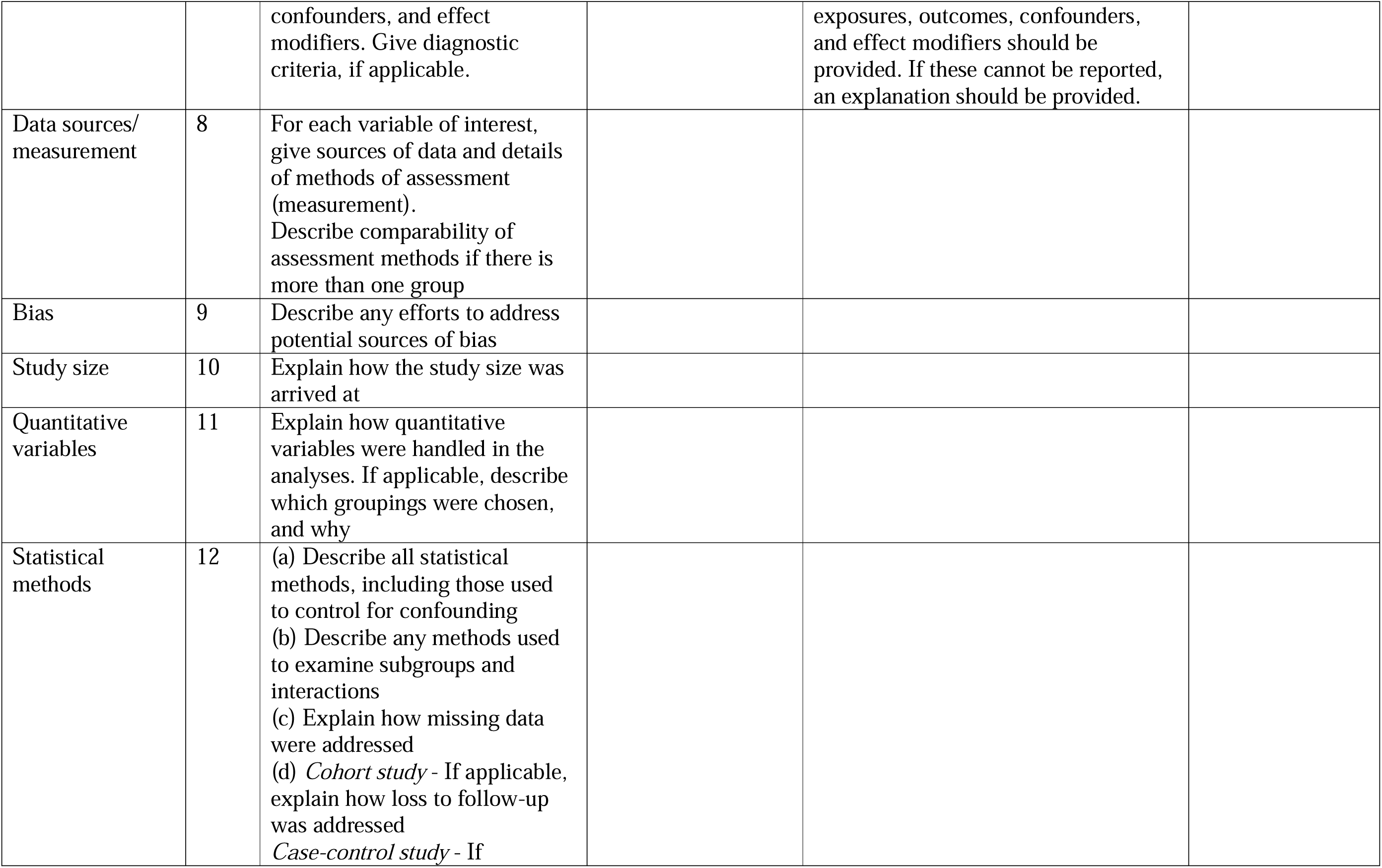
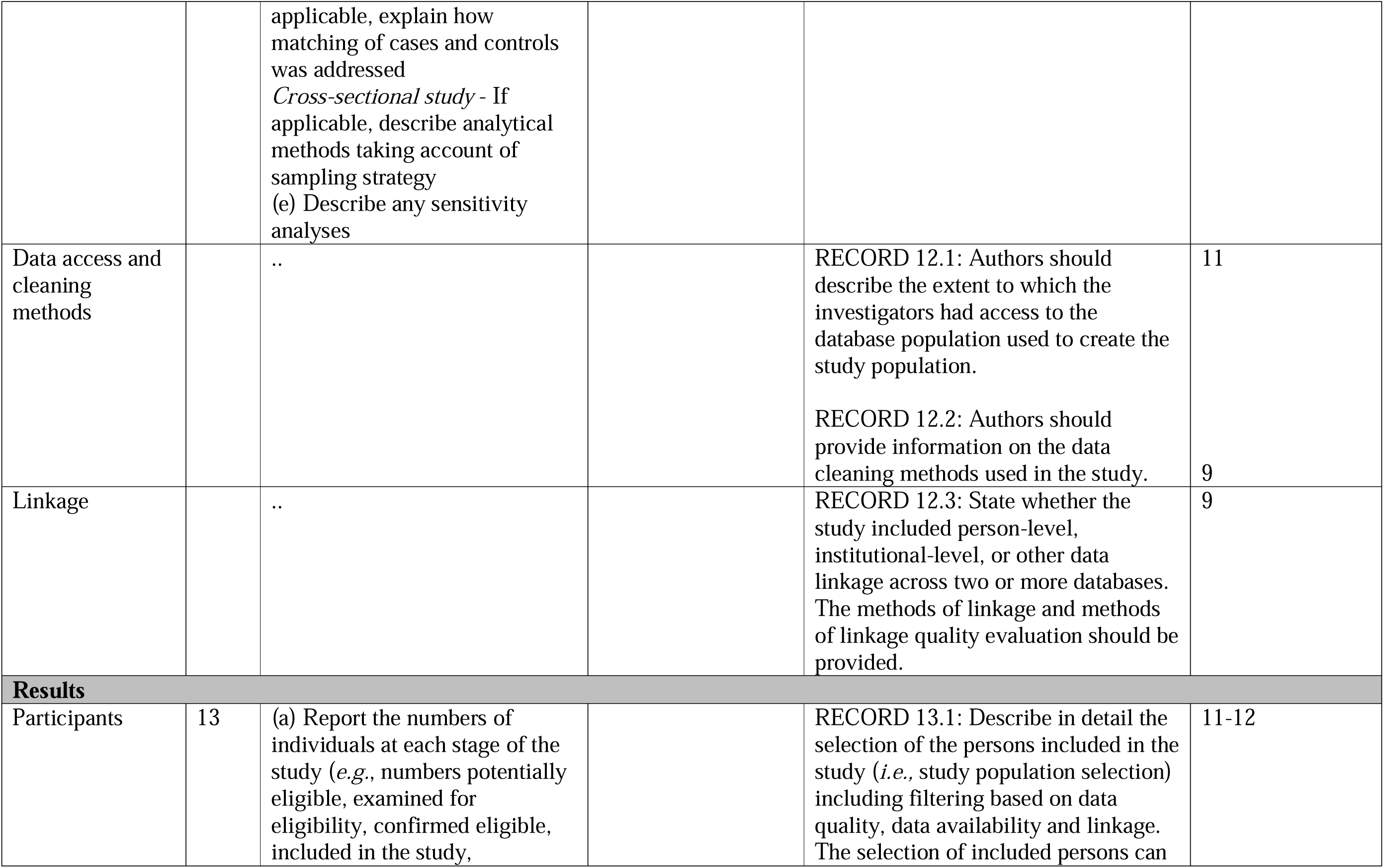
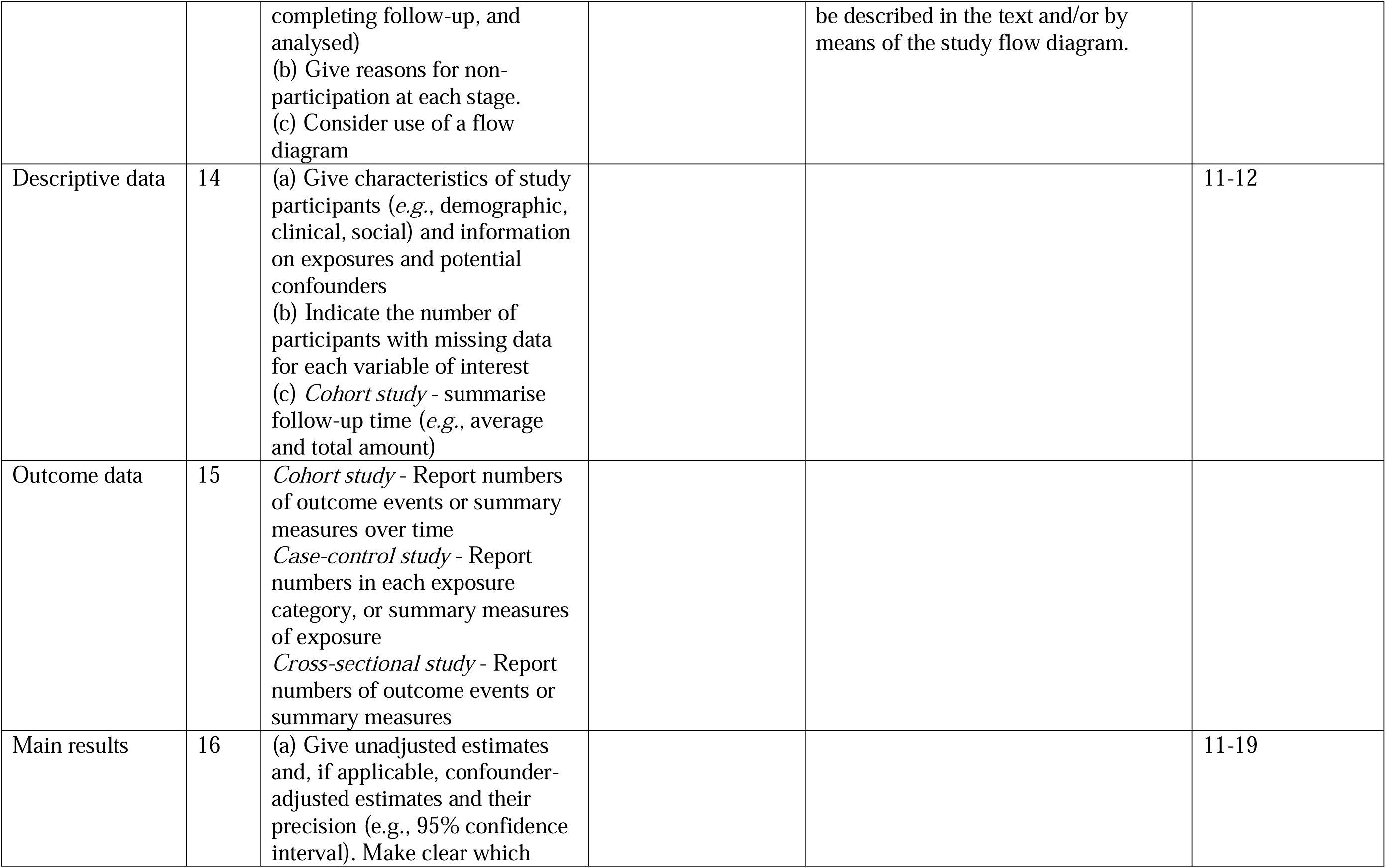
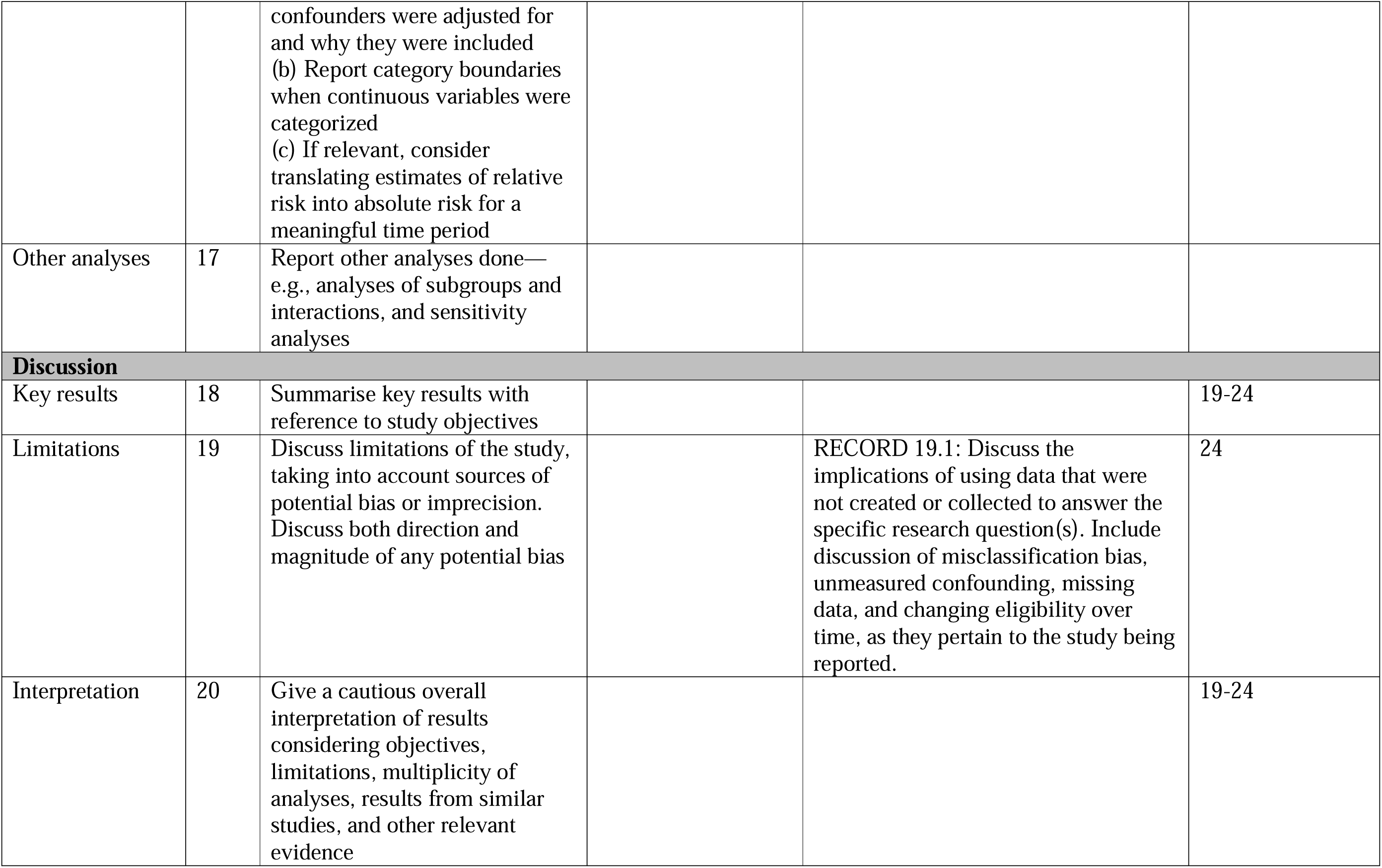
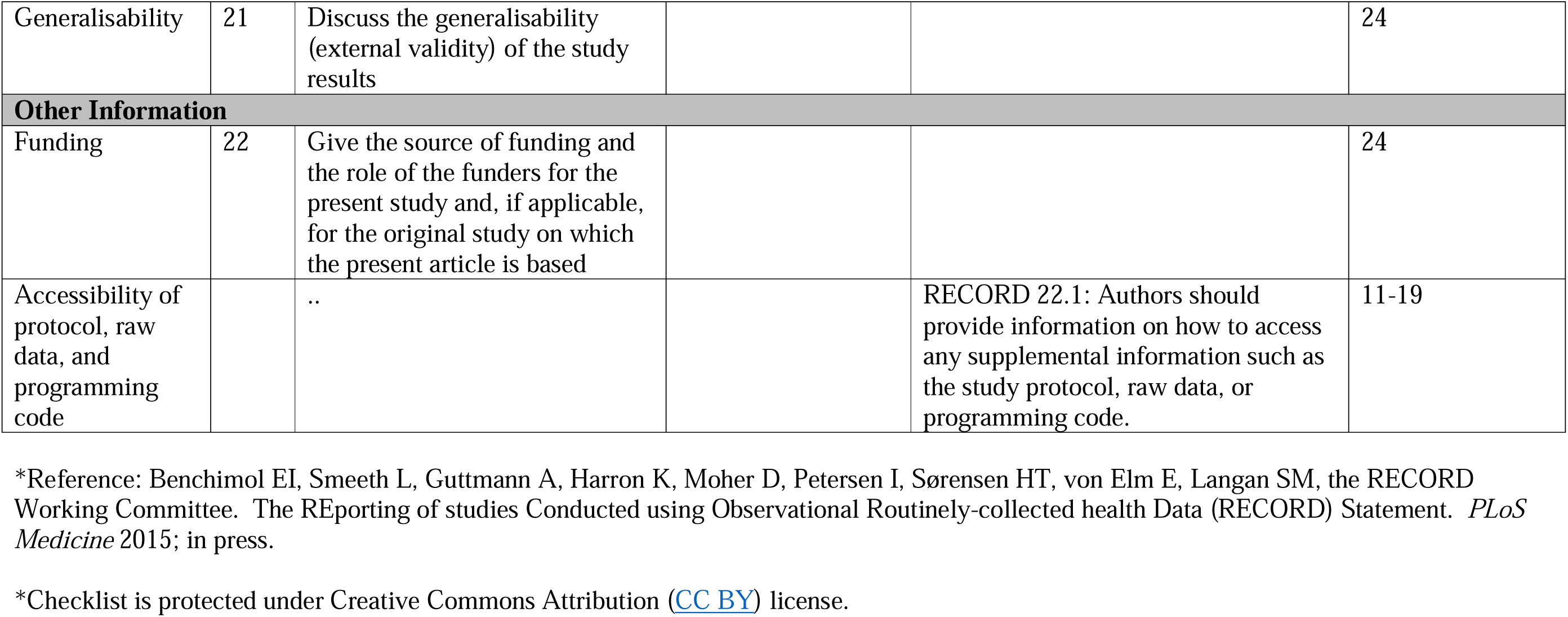

|  | **Item No.** | **STROBE items** | **Location in manuscript where items are reported** | **RECORD items** | **Location in manuscript where items are reported** |
| --- | --- | --- | --- | --- | --- |
| **Title and abstract** |  |  |  |  |  |
|  | 1 | (a) Indicate the study's design with a commonly used term in the title or the abstract (b) Provide in the abstract an informative and balanced summary of what was done and what was found |  | RECORD 1.1: The type of data used should be specified in the title or abstract. When possible, the name of the databases used should be included.<br>RECORD 1.2: If applicable, the geographic region and timeframe within which the study took place should be reported in the title or abstract.<br>RECORD 1.3: If linkage between databases was conducted for the study, this should be clearly stated in the title or abstract. | 1<br>1<br>1 |
| **Introduction** |  |  |  |  |  |
| Background rationale | 2 | Explain the scientific background and rationale for the investigation being reported |  |  | 3-5 |
| Objectives | 3 | State specific objectives, including any prespecified hypotheses |  |  | 5 |
| **Methods** |  |  |  |  |  |
| Study Design | 4 | Present key elements of study design early in the paper |  |  | 5 |
| Setting | 5 | Describe the setting, locations, and relevant dates, including periods of recruitment, exposure, follow-up, and data collection |  |  | 5-6 |
| Participants | 6 | (a) *Cohort study* - Give the eligibility criteria, and the sources and methods of selection of participants. Describe methods of follow-up<br>*Case-control study* - Give the eligibility criteria, and the sources and methods of case ascertainment and control selection. Give the rationale for the choice of cases and controls<br>*Cross-sectional study* - Give the eligibility criteria, and the sources and methods of selection of participants<br>(b) *Cohort study* - For matched studies, give matching criteria and number of exposed and unexposed<br>*Case-control study* - For matched studies, give matching criteria and the number of controls per case |  | RECORD 6.1: The methods of study population selection (such as codes or algorithms used to identify subjects) should be listed in detail. If this is not possible, an explanation should be provided.<br>RECORD 6.2: Any validation studies of the codes or algorithms used to select the population should be referenced. If validation was conducted for this study and not published elsewhere, detailed methods and results should be provided.<br>RECORD 6.3: If the study involved linkage of databases, consider use of a flow diagram or other graphical display to demonstrate the data linkage process, including the number of individuals with linked data at each stage. | 6 & 9<br>6<br>9 |
| Variables | 7 | Clearly define all outcomes, exposures, predictors, potential |  | RECORD 7.1: A complete list of codes and algorithms used to classify | 9 |
|  |  | confounders, and effect modifiers. Give diagnostic criteria, if applicable. |  | exposures, outcomes, confounders, and effect modifiers should be provided. If these cannot be reported, an explanation should be provided. |  |
| Data sources/<br>measurement | 8 | For each variable of interest, give sources of data and details of methods of assessment (measurement). Describe comparability of assessment methods if there is more than one group |  |  |  |
| Bias | 9 | Describe any efforts to address potential sources of bias |  |  |  |
| Study size | 10 | Explain how the study size was arrived at |  |  |  |
| Quantitative<br>variables | 11 | Explain how quantitative variables were handled in the analyses. If applicable, describe which groupings were chosen, and why |  |  |  |
| Statistical<br>methods | 12 | (a) Describe all statistical methods, including those used to control for confounding<br>(b) Describe any methods used to examine subgroups and interactions<br>(c) Explain how missing data were addressed<br>(d) *Cohort study* - If applicable, explain how loss to follow-up was addressed<br>*Case-control study* - If applicable, explain how matching of cases and controls was addressed<br>*Cross-sectional study* - If applicable, describe analytical methods taking account of sampling strategy<br>(e) Describe any sensitivity analyses |  |  |  |
| Data access and cleaning methods |  | .. |  | RECORD 12.1: Authors should describe the extent to which the investigators had access to the database population used to create the study population.<br>RECORD 12.2: Authors should provide information on the data cleaning methods used in the study. | 11<br>9 |
| Linkage |  | .. |  | RECORD 12.3: State whether the study included person-level, institutional-level, or other data linkage across two or more databases. The methods of linkage and methods of linkage quality evaluation should be provided. | 9 |
| **Results** |  |  |  |  |  |
| Participants | 13 | (a) Report the numbers of individuals at each stage of the study (*e.g.*, numbers potentially eligible, examined for eligibility, confirmed eligible, included in the study, |  | RECORD 13.1: Describe in detail the selection of the persons included in the study (*i.e.*, study population selection) including filtering based on data quality, data availability and linkage. The selection of included persons can | 11-12 |
|  |  | completing follow-up, and analysed)<br>(b) Give reasons for non-participation at each stage.<br>(c) Consider use of a flow diagram |  | be described in the text and/or by means of the study flow diagram. |  |
| Descriptive data | 14 | (a) Give characteristics of study participants (*e.g.*, demographic, clinical, social) and information on exposures and potential confounders<br>(b) Indicate the number of participants with missing data for each variable of interest<br>(c) *Cohort study* - summarise follow-up time (*e.g.*, average and total amount) |  |  | 11-12 |
| Outcome data | 15 | *Cohort study* - Report numbers of outcome events or summary measures over time<br>*Case-control study* - Report numbers in each exposure category, or summary measures of exposure<br>*Cross-sectional study* - Report numbers of outcome events or summary measures |  |  |  |
| Main results | 16 | (a) Give unadjusted estimates and, if applicable, confounder-adjusted estimates and their precision (*e.g.*, 95% confidence interval). Make clear which<br>confounders were adjusted for and why they were included<br>(b) Report category boundaries when continuous variables were categorized<br>(c) If relevant, consider translating estimates of relative risk into absolute risk for a meaningful time period |  |  | 11-19 |
| Other analyses | 17 | Report other analyses done—e.g., analyses of subgroups and interactions, and sensitivity analyses |  |  |  |
| **Discussion** |  |  |  |  |  |
| Key results | 18 | Summarise key results with reference to study objectives |  |  | 19-24 |
| Limitations | 19 | Discuss limitations of the study, taking into account sources of potential bias or imprecision. Discuss both direction and magnitude of any potential bias |  | **RECORD 19.1:** Discuss the implications of using data that were not created or collected to answer the specific research question(s). Include discussion of misclassification bias, unmeasured confounding, missing data, and changing eligibility over time, as they pertain to the study being reported. | 24 |
| Interpretation | 20 | Give a cautious overall interpretation of results considering objectives, limitations, multiplicity of analyses, results from similar studies, and other relevant evidence |  |  | 19-24 |
| Generalisability | 21 | Discuss the generalisability (external validity) of the study results |  |  | 24 |
| **Other Information** |  |  |  |  |  |
| Funding | 22 | Give the source of funding and the role of the funders for the present study and, if applicable, for the original study on which the present article is based |  |  | 24 |
| Accessibility of protocol, raw data, and programming code |  | .. |  | RECORD 22.1: Authors should provide information on how to access any supplemental information such as the study protocol, raw data, or programming code. | 11-19 |

\*Reference: Benchimol EI, Smeeth L, Guttman A, Harron K, Moher D, Petersen I, Sørensen HT, von Elm E, Langan SM, the RECORD Working Committee. The REporting of studies Conducted using Observational Routinely-collected health Data (RECORD) Statement. *PLoS Medicine* 2015; in press.
\*Checklist is protected under Creative Commons Attribution ([CC BY](https://creativecommons.org/licenses/by/4.0/)) license.

## REFERENCES

1. World Health Organization. Ethics and governance of artificial intelligence for health. Guidance on large multi-modal models [Internet]. Geneva: World Health Organization; 2024 [cited 2026 Mar 7]. 1–98 p. Available from: https://iris.who.int/server/api/core/bitstreams/e9e62c65-6045-481e-bd04-20e206bc5039/content

2. Wiens J, Saria S, Sendak M, Ghassemi M, Liu VX, Doshi-Velez F, et al. Do no harm: a roadmap for responsible machine learning for health care. Nat Med. 2019 Sep;25(9):1337–40. doi:10.1038/s41591-019-0609-x PubMed PMID: 31537911.

3. Topol EJ. High-performance medicine: the convergence of human and artificial intelligence. Nat Med. 2019 Jan 7;25(1):44–56. doi:10.1038/s41591-018-0300-7 PubMed PMID: 30617339.

4. Wahl B, Cossy-Gantner A, Germann S, Schwalbe NR. Artificial intelligence (AI) and global health: how can AI contribute to health in resource-poor settings? BMJ Glob Health. 2018 Aug 29;3(4):e000798. doi:10.1136/bmjgh-2018-000798 PubMed PMID: 10.1136/bmjgh-2018-000798.

5. Kelly CJ, Karthikesalingam A, Suleyman M, Corrado G, King D. Key challenges for delivering clinical impact with artificial intelligence. BMC Med. 2019 Oct 29;17(1):195. doi:10.1186/s12916-019-1426-2 PubMed PMID: 31665002.

6. Nagendran M, Chen Y, Lovejoy CA, Gordon AC, Komorowski M, Harvey H, et al. Artificial intelligence versus clinicians: systematic review of design, reporting standards, and claims of deep learning studies. BMJ. 2020 Mar 25;368:m689. doi:10.1136/bmj.m689 PubMed PMID: 32213531.

7. Ioannidis JPA. Why Most Published Research Findings Are False. PLoS Med. 2005 Aug 30;2(8):e124. doi:10.1371/journal.pmed.0020124 PubMed PMID: 16060722.

8. Sculley D, Holt G, Golovin D, Davydov E, Phillips T, Ebner D, et al. Hidden Technical Debt in Machine Learning Systems. Adv Neural Inf Process Syst [Internet]. 2015 [cited 2026 Mar 21];28:1–9. Available from: https://proceedings.neurips.cc/paper_files/paper/2015/file/86df7dcfd896fcaf2674f757a2463eba-Paper.pdf

9. Turner EH, Matthews AM, Linardatos E, Tell RA, Rosenthal R. Selective publication of antidepressant trials and its influence on apparent efficacy. N Engl J Med. 2008 Jan 17;358(3):252–60. doi:10.1056/nejmsa065779 PubMed PMID: 18199864.

10. Lexchin J, Bero LA, Djulbegovic B, Clark O. Pharmaceutical industry sponsorship and research outcome and quality: systematic review. BMJ. 2003 May 31;326(7400):1167–70. doi:10.1136/bmj.326.7400.1167 PubMed PMID: 12775614.

11. World Bank Group. The World Bank Press Release [Internet]. 2023 [cited 2026 Mar 21]. Investments in Digital Can Accelerate Improvements in Health Care. Available from: https://www.worldbank.org/en/news/press-release/2023/08/19/investments-in-digital-can-accelerate-improvements-in-health-care

12. World Economic Forum. Health and Healthcare Systems [Internet]. 2024 [cited 2026 Mar 21]. Billions of dollars have been invested in healthcare AI. But are we spending in the right places? . Available from: https://www.weforum.org/stories/2024/11/healthcare-health-ai/

13. Aju OG, Osubor VI. Accent-Based Speech Recognition Model for the Major Nigerian Ethnics English Speakers. FUOYE Journal of Pure and Applied Sciences . 2024 Dec 25;9(3):67–82. doi:10.55518/fjpas.FKSP5386

14. Mekniran W, Diethelm W, Stalder V, Fleisch E, Kowatsch T, Jovanova M. Digital health technologies and stakeholder incentives in type-2 diabetes prevention. Digit Health. 2026 Jan 1;12:20552076261425400. doi:10.1177/20552076261425402 PubMed PMID: 41836627.

15. Liu X, Faes L, Kale AU, Wagner SK, Fu DJ, Bruynseels A, et al. A comparison of deep learning performance against health-care professionals in detecting diseases from medical imaging: a systematic review and meta-analysis. Lancet Digit Health. 2019 Oct 1;1(6):e271–97. doi:10.1016/S2589-7500(19)30123-2 PubMed PMID: 33323251.

16. Liu X, Cruz Rivera S, Moher D, Calvert MJ, Denniston AK, Ashrafian H, et al. Reporting guidelines for clinical trial reports for interventions involving artificial intelligence: the CONSORT-AI extension. Lancet Digit Health. 2020 Oct 1;2(10):e537–48. doi:10.1016/S2589-7500(20)30218-1 PubMed PMID: 33328048.

17. Benchimol EI, Smeeth L, Guttmann A, Harron K, Moher D, Peteresen I, et al. The REporting of studies Conducted using Observational Routinely-collected health Data (RECORD) Statement. PLoS Med. 2015;12(10):e1001885. doi:10.1371/journal.pmed.1001885 PubMed PMID: 26440803.

18. Wynants L, Van Calster B, Collins GS, Riley RD, Heinze G, Schuit E, et al. Prediction models for diagnosis and prognosis of covid-19: systematic review and critical appraisal. BMJ. 2020 Apr 7;369:m1328. doi:10.1136/bmj.m1328 PubMed PMID: 32265220.

19. US Food & Drug Administration. US F&DA Digital Center of Excellence [Internet]. 2025 [cited 2026 Mar 21]. Request For Public Comment: Measuring and Evaluating Artificial Intelligence-enabled Medical Device Performance in the Real-World. Available from: https://www.fda.gov/medical-devices/digital-health-center-excellence/request-public-comment-measuring-and-evaluating-artificial-intelligence-enabled-medical-device

20. Windecker D, Baj G, Shiri I, Kazaj PM, Kaesmacher J, Gräni C, et al. Generalizability of FDA-Approved AI-Enabled Medical Devices for Clinical Use. JAMA Netw Open. 2025 Apr 1;8(4):e258052–e258052. doi:10.1001/jamanetworkopen.2025.8052 PubMed PMID: 40305017.

21. Akpan F. Bridging Nigeria’s Healthcare Gaps through Artificial Intelligence. News Agency of Nigeria [Internet]. 2026 Jan 15 [cited 2026 Mar 21]. Available from: https://nannews.ng/bridging-nigerias-healthcare-gaps-through-artificial-intelligence/

22. Celi LA, Cellini J, Charpignon ML, Dee EC, Dernoncourt F, Eber R, et al. Sources of bias in artificial intelligence that perpetuate healthcare disparities—A global review. PLOS Digital Health. 2022 Mar 1;1(3):e0000022. doi:10.1371/journal.pdig.0000022

23. Finlayson SG, Subbaswamy A, Singh K, Bowers J, Kupke A, Zittrain J, et al. The Clinician and Dataset Shift in Artificial Intelligence. N Engl J Med. 2021 Jul 15;385(3):283–6. doi:10.1056/nejmc2104626 PubMed PMID: 34260843.

24. Beede E, Baylor E, Hersch F, Iurchenko A, Wilcox L, Ruamviboonsuk P, et al. A Human-Centered Evaluation of a Deep Learning System Deployed in Clinics for the Detection of Diabetic Retinopathy. Conference on Human Factors in Computing Systems - Proceedings. 2020 Apr 21;1–12. doi:10.1145/3313831.3376718

25. Obermeyer Z, Powers B, Vogeli C, Mullainathan S. Dissecting racial bias in an algorithm used to manage the health of populations. Science. 2019 Oct 25;366(6464):447–53. doi:10.1126/science.aax2342 PubMed PMID: 31649194.

26. Seyyed-Kalantari L, Zhang H, McDermott MBA, Chen IY, Ghassemi M. Underdiagnosis bias of artificial intelligence algorithms applied to chest radiographs in under-served patient populations. Nat Med. 2021 Dec 1;27(12):2176–82. doi:10.1038/s41591-021-01595-0 PubMed PMID: 34893776.

27. Larrazabal AJ, Nieto N, Peterson V, Milone DH, Ferrante E. Gender imbalance in medical imaging datasets produces biased classifiers for computer-aided diagnosis. Proc Natl Acad Sci U S A. 2020 Jun 9;117(23):12592–4. doi:10.1073/pnas.1919012117 PubMed PMID: 32457147.

28. Keshavarz MH. An investigation into pronunciation problems of Hausa-speaking learners of English. Int Online J Educ Teach [Internet]. 2017 [cited 2026 Mar 21];4(1):61–72. Available from: https://iojet.org/index.php/IOJET/article/view/152/150

29. Breuninger M, Van Ginneken B, Philipsen RHHM, Mhimbira F, Hella JJ, Lwilla F, et al. Diagnostic Accuracy of Computer-Aided Detection of Pulmonary Tuberculosis in Chest Radiographs: A Validation Study from Sub-Saharan Africa. PLoS One. 2014 Sep 5;9(9):e106381. doi:10.1371/journal.pone.0106381 PubMed PMID: 25192172.

30. Qin ZZ, Van der Walt M, Moyo S, Ismail F, Maribe P, Denkinger CM, et al. Computer-aided detection of tuberculosis from chest radiographs in a tuberculosis prevalence survey in South Africa: external validation and modelled impacts of commercially available artificial intelligence software. Lancet Digit Health. 2024 Sep 1;6(9):e605–13. doi:10.1016/S2589-7500(24)00118-3 PubMed PMID: 39033067.

31. Coiera E, Ash J, Berg M. The Unintended Consequences of Health Information Technology Revisited. Yearb Med Inform. 2016 Nov 10;10(1):169. doi:10.15265/iy-2016-014 PubMed PMID: 27830246.

